# IL-1 and IL-6 inhibitor hypersensitivity link to common HLA-DRB1*15 alleles

**DOI:** 10.1101/2020.08.10.20172338

**Authors:** Jill A. Hollenbach, Michael J. Ombrello, Adriana H. Tremoulet, Gonzalo Montero-Martin, Sampath Prahalad, Scott Canna, Chisato Shimizu, Gail Deutsch, Serena Tan, Elaine F. Remmers, Dimitri Monos, Omkar K. Phadke, Jianpeng Xu, Jaime S. Rosa Duque, Gilbert T. Chua, Vamsee Mallajosyula, Debopam Ghosh, Ann Marie Szymanski, Danielle Rubin, Jane C. Burns, Marcelo A. Fernandez-Vina, Elizabeth D. Mellins, Vivian E. Saper, the Drug Hypersensitivity Consortium

## Abstract

**Background:** Drug reaction with eosinophilia and systemic symptoms (DRESS), a severe, delayed hypersensitivity reaction (DHR), is underrecognized, particularly in patients with inflammatory diseases. Atypical features arising during interleukin 1 (IL-1) and interleukin 6 (IL-6) inhibitor treatment of systemic onset juvenile arthritis (sJIA) and adult onset Stills disease (AOSD) suggested this serious DHR.

**Methods:** We conducted a multi-center, retrospective study of a convenience sample of IL-1 or IL-6 inhibitor-exposed cases of sJIA, sJIA-like disease, AOSD, and Kawasaki disease (n=105 participants with sequence data or samples for HLA typing). Using pre-determined parameters, including RegiSCAR scoring for DRESS, subjects were classified as DHR-positive or drug-tolerant. HLA allele frequency was compared in cases versus drug-tolerant controls and versus the largest available sJIA cohort, collected by the International Childhood Arthritis Genetics Consortium (INCHARGE).

**Results:** DHR features included eosinophilia (94%), AST-ALT elevation (70%), and non-evanescent rash (94%; 82% involving face). Cytokine storm syndrome during inhibitor treatment, a known complication of DRESS, was frequent (67%). A striking enrichment for common HLA-DRB1*15 haplotypes was present in DHR cases relative to drug-tolerant controls and was corroborated with the INCHARGE sJIA cohort (p=1×10^−10^). This association differs from the HLA-DR association with sJIA itself and is far stronger, as expected for HLA-associated DHR risk.

**Conclusions:** Serious, at times fatal, DRESS-type reactions occur in a subset of patients exposed to IL-1/IL-6 inhibitors and strongly associate with HLA-DRB1*15 alleles that are common across ancestries. HLA testing before prescription warrants consideration, and alertness to DHR during treatment across conditions is prudent to improve safety.

## Introduction

Severe, potentially fatal, delayed hypersensitivity reactions (DHR) are underrecognized due to their complexity and variable presentation.^1,2^ Particularly during treatment of inflammatory illnesses, DHR may be misinterpreted as disease flares. The most serious types of DHR classify as severe cutaneous adverse reactions (SCARs). Among these is drug reaction with eosinophilia and systemic symptoms (DRESS). Typical features of DRESS are latency (days to months) after drug initiation, fever, extensive rash, hematologic manifestations (eosinophilia and atypical lymphocytosis), involvement of various deep organs, and often an extended time to recovery, even after the offending drug is stopped. Prevalence of DRESS in the United States is estimated at 2.2 cases per 100,00 patients^3^, a low rate among drug reaction types. Recognition of these serious, less common drug reactions is both imperative and challenging.

Increasingly, pharmacogenetic data link drug-specific reaction risk with particular human leukocyte antigen (HLA) class I and/or class II alleles. HLA alleles exhibit by far the most robust association with severe DHR among genetic factors. Moreover, these HLA associations have proven to be substantially stronger with much higher odds ratios and more complete penetrance than most of the well-known HLA allelic associations with autoimmune diseases.^4,5^ In addition to providing clues to pathogenesis, the finding of an HLA/DHR association allows preventative HLA screening pre-prescription. Some well-characterized HLA associations are specific to alleles found primarily in particular populations; others have been linked to relatively common alleles with a wide global distribution. The cost/benefit ratio of HLA screening to prevent DHR in at-risk individuals improves as the population frequency of the HLA risk allele increases.^5,6^

HLA molecules function to present peptides to T cells through binding to T cell surface receptors for antigen. In some DHR, the offending drugs have been shown to interact directly with HLA molecules, which in turn stimulate T cell responses; the drug interaction can alter the repertoire of peptides bound to HLA.^5^ Thus, HLA associations with DHR implicate T cells as immune effectors. This implication is corroborated by evidence from biopsies of DHR-associated skin rashes, which show infiltration of activated T cells.^7^

Evidence suggests that underlying immune dysregulation greatly enhances the chance that a T cell response to an offending drug will escalate to clinical significance.^8^ Consistent with this, we observed DRESS in systemic juvenile idiopathic arthritis (sJIA), a chronic inflammatory disease of childhood with unknown etiology.^9^ We noted DRESS among sJIA patients who developed an unusual, non-infectious parenchymal diffuse lung disease (DLD) during immunosuppressive treatment with inhibitors of IL-1 (anakinra, canakinumab, rilonacept) or of IL-6 (tocilizumab).^10^ We hypothesized that DHR to these drugs was underrecognized in sJIA and its adult counterpart, adult onset Still’s disease (AOSD).^11^ Through a multicenter effort, we identified sJIA, sJIA-like disease and AOSD patients with possible inhibitor-triggered DHR, with and without lung disease (n=56), or with apparent drug tolerance for >1 year (n=30). Consistent with the rising use of the inhibitors in sJIA treatment, 66% of DHR cases began between 2015 and the end of 2020. Subjects in this convenience cohort were required to have genome-wide sequence data or samples for HLA typing. Also included in this study was a small cohort of Kawasaki disease (KD) patients (n=19) in a phase I/IIa trial of anakinra (NCT-02179853)^12^; all had samples for HLA genotyping. Based on clinical data, we classified subjects as having DHR, suspected anakinra reaction (sAR) or no drug reaction (i.e., drug-tolerant) (**figure 1**).^13^ We compared the DHR/sAR cases to the drug-tolerant controls for demographics and clinical features. To investigate genetic risk of DHR/sAR, we carried out HLA association analysis. We increased the size of our sJIA control group for this analysis by accessing HLA data from the INCHARGE sJIA case-control collection, which includes 773 sJIA cases.^14^

**Figure 1:**
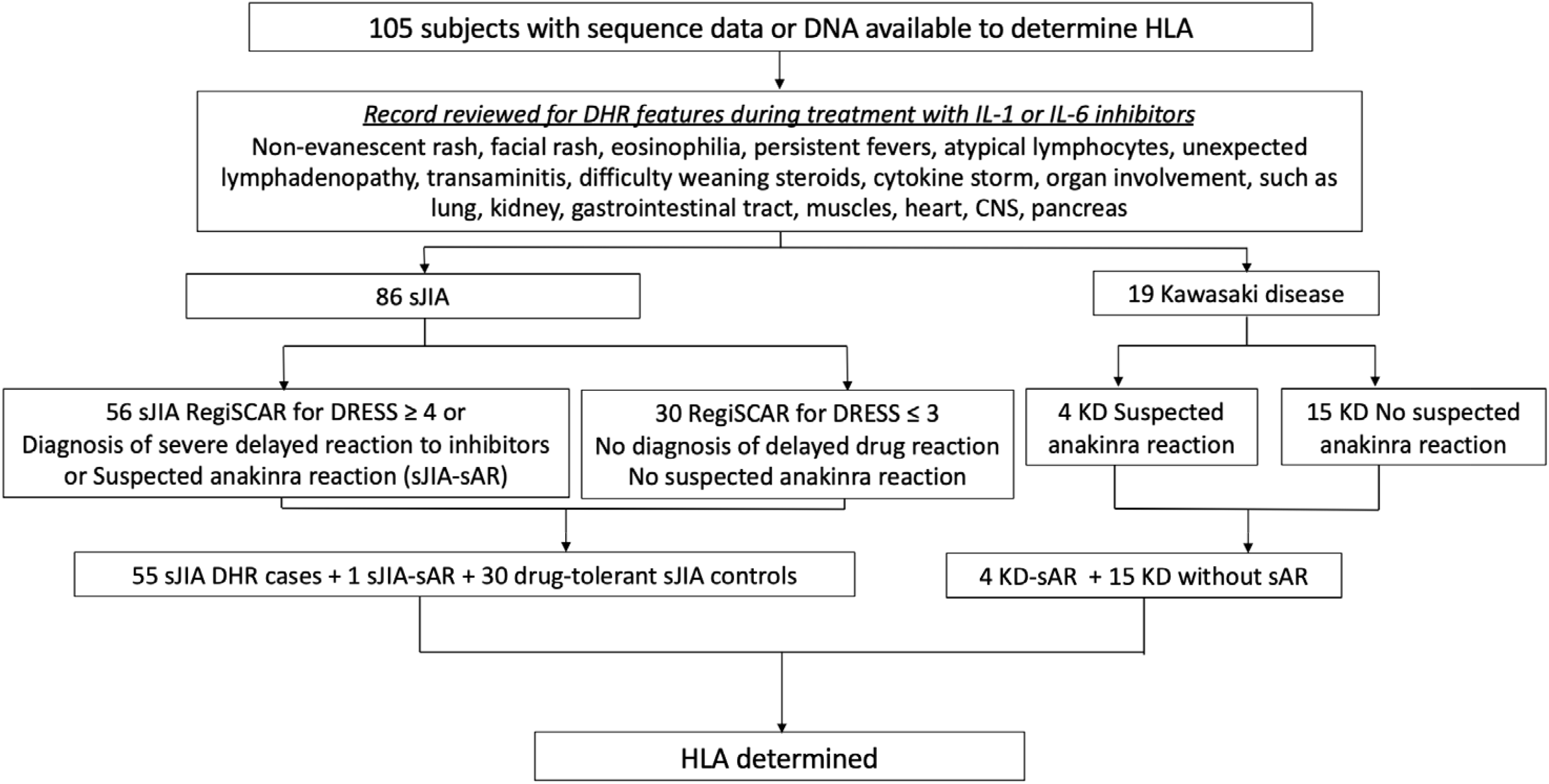
Study design. RegiSCAR for DRESS scoring was performed as described.^12^ HLA genotyping was performed as described in Methods.

## Methods

### Approvals

This study was approved by Institutional Review Board (IRB), Stanford University, and by local IRB from contributing institutions, based on local practice. Informed consent was obtained for HLA genotyping or sharing of genetic data.

### Subjects

Subjects were identified through outreach to clinicians at 35 centers in the US and Canada. All subjects were exposed to anakinra, canakinumab, rilonacept or tocilizumab and had available either whole exome sequence data or a sample for HLA typing.

### Clinical data collection and analysis

Clinical information was collected into a privacy-protected electronic database or by direct communication with the physician case reporter. DHR diagnosis was based on assessment by treating physician or on scoring as definite or probable DRESS, using the validated RegiSCAR system.^13^ Suspected anakinra reaction (sAR) required an AEC ≥500. sJIA-sAR required >2 occurrences of unexplained eosinophilia during treatment. KD-sAR required an increase in eosinophils of ≥50% over pre-treatment value at study baseline. Data for full RegiSCAR scoring of KD subjects were unavailable. Drug-tolerance (sJIA/AOSD) was defined as no drug reaction during ≥1 year of IL-1 or IL-6 inhibitor treatment with discontinuation of steroids or ≥6wks of steroid at <0.2 mg/kg/day. Presumed drug-tolerance (KD) was defined as absence of eosinophilia during anakinra exposure (9-46 days). sJIA and KD subjects were classified (cases/controls) by a board-certified allergist (VS), prior to HLA determination (**figure 1**). Ancestry, demographic and clinical data are on **tables S1a-b, S2a-b, S3**.

### HLA association analysis

As described in appendix, genomic DNA was extracted from blood or tissue, and HLA genotyping was performed by one of several methods. To examine the *a priori* hypothesis that HLA-DRB1*15:01 is associated with DHR in drug-treated patients, a 2 x 2 contingency table using Fisher’s exact test was employed. Analyses assumed the dominant model and were conducted in the R environment for statistical computing^15^ and p-values and odds ratios with 95% confidence intervals were obtained using the ‘oddsratio.fisher’ function in the ‘epitools’ package.^16^

Classical HLA alleles of the sJIA-DHR patients were also compared to those of the INCHARGE cohort, the largest available sJIA collection.^14^ INCHARGE sJIA cases and healthy controls matching the genetic ancestry of 24 sJIA-DHR subjects with WES data were identified by principal component analysis (PCA). PCA yielded a tight cluster of INCHARGE sJIA cases (n=550) and control subjects (n=3279), together with every subject of self-reported White (European) ancestry analyzed by PCA (n=14) from the sJIA-DHR cohort (**figure S1**). As self-reported White ancestry showed perfect concordance with PCA cluster membership, 15 self-reported White sJIA-DHR subjects without WES were added as cases. Genomic control inflation factors (λ_GC_) were 1.00-1.05, demonstrating robust matching. Association testing was performed as above (**tables 1,2**) and by logistic regression (**table S4**), as described^21^. Additional information on all method sections are in appendix.

**Table 1:** HLA class II allele association with hypersensitivity to IL-1 and IL-6 inhibitors. Within-group analyses for ancestries other than White/European were not done due to insufficient numbers and lack of matching of individual groups among the “all” group for sJIA/INCHARGE sJIA.

| HLA allele | Ancestry | Cases | Controls |  |  |  |
| --- | --- | --- | --- | --- | --- | --- |
| sJIA |  | sJIA-[DHR+sAR] | Drug-tolerant sJIA | INCHARGE sJIA | P value | OR (95% CI) |
| HLA-DRB1*15:01 | self-ID White | 24/29 (83%) | 0/19 (0%) |  | 7x10<sup>-9</sup> | Inf (14.49-Infinite) |
|  | self-ID White vs European | 24/29 (83%) |  | 130/550 (24%) | 1x10<sup>-10</sup> | 15.4 (5.6-52.8) |
|  | European vs European | 13/14 (93%) |  | 130/550 (24%) | 1x10<sup>-7</sup> | 41.7 (6.2-1772.3) |
| HLA-DRB1*15:01 | self-ID non-White | 15/27 (56%) | 0/11 (0%) |  |  |  |
| HLA-DRB1*15:XX | self-ID non-White | 20/27 (74%) | 2/11 (18%) |  |  |  |
|  | All | 44/56 (79%) |  | 182/773 (24%) |  |  |
| Kawasaki disease |  | KD only | Drug-tolerant KD |  |  |  |
| HLA-DRB1*15:01 | All | 2/4 (50%) | 0/15 (0%) |  |  |  |
| HLA-DRB1*15:XX | All | 3/4 (75%) | 2/15 (13%)<sup>1</sup> |  |  |  |
| sJIA + Kawasaki disease |  | sJIA-[DHR+sAR]+KD | Drug-tolerant sJIA+KD |  |  |  |
| HLA-DRB1*15:01 | All | 41/60 (68%) | 0/45 (0%) |  |  |  |
| HLA-DRB1*15:XX | All | 47/60 (78%) | 4/45 (9%) |  |  |  |
| HLA-DQB1*06:02 | All | 43/60 (72%) | 3/45 (7%) |  |  |  |

DHR+sAR, Delayed hypersensitivity + suspected anakinra reaction; sJIA, systemic juvenile idiopathic arthritis; INCHARGE, International Childhood Arthritis Genetics Consortium; OR (95% CI), Odds ratio and 95% confidence interval; European: sJIA DHR+sAR cases were ancestry-matched by PCA to the sJIA INCHARGE cohort (figure s1); Inf, infinite; KD, Kawasaki disease; HLA-DRB1\*15:XX: All HLA-DRB1\*15 alleles
<sup>1</sup>HLA-DRB1\*15:02 in two individuals, treated briefly (12 and 28 days) with anakinra

**Table 2:** HLA-DRB1*11:01 is sJIA-associated in the sJIA-[DHR+sAR] cohort.

| HLA allele | Ancestry | sJIA-[DHR+sAR] | INCHARGE sJIA | INCHARGE controls | *P* value | OR (95% CI) |
| --- | --- | --- | --- | --- | --- | --- |
| HLA-DRB1*11:01 | European vs<br>European INCHARGE | 4/14 (29%) | 103/550 (19%) |  | NS |  |
|  |  |  |  | 313/3279 (10%) | 0.04 | 3.8 (.86-13.2) |
| HLA-DRB1*11:01 | Self-ID White vs<br>European INCHARGE | 7/29 (24%) | 103/550 (19%) |  | NS |  |
|  |  |  |  | 313/3279 (10%) | 0.02 | 3.0 (1.1-7.4)<sup>1</sup> |
| HLA-DRB1*15:01 |  |  | 130/550 (24%) | 822/3279 (25%) | NS |  |
| HLA-DRB1*11:01 | All vs<br>All INCHARGE | 12/56 (21%) | 160/773 (21%) |  |  |  |
|  |  |  |  | 649/6812 (10%) |  |  |
| HLA-DRB1*15:XX |  |  | 182/773 (24%) | 1736/6812 (25%) | NS |  |

OR (95% CI), Odds ratio and 95% confidence interval; NS, not significant; sJIA, systemic juvenile idiopathic arthritis; DHR+sAR, Delayed hypersensitivity plus suspected anakinra reaction; INCHARGE, International Childhood Arthritis Genetics Consortium; European, sJIA DHR+sAR cases were ancestry-matched by PCA to the sJIA INCHARGE; HLA-DRB1\*15:XX: All HLA-DRB1\*15 alleles
<sup>1</sup>Results are consistent with published data from INCHARGE consortium study of HLA association with sJIA.<sup>21</sup>
Odds ratios are only shown for significant associations. Between group analyses using the full DHR and INCHARGE cohorts were not performed due to lack of ancestral matching. Comparisons with the INCHARGE cohorts using logistic regression are provided on table S4.

## Results

### DHR, often unrecognized, occurs in a subset of sJIA/AOSD patients treated with IL-1 or IL-6 inhibitors

To collect cases for an analysis of DHR to IL-1 inhibitors (anakinra, canakinumab, rilonacept) or an IL-6 inhibitor (tocilizumab) in sJIA and AOSD, we sought patients with diffuse lung disease (DLD; see **figure 2**), as we suspected this group included patients with DHR. We also sought patients with DHR only and drug-tolerant controls. We confirmed classification for DHR or drug tolerance using specified criteria (see Methods and **figure 1**). All but 1 of the sJIA/AOSD patients with DHR were classified as drug-related eosinophilia with systemic symptoms (DRESS); the single exception was classified as suspected anakinra reaction (sAR). The DHR group and the drug-tolerant group were similar for age of onset of sJIA, male: female ratio, and range of ethnicities (**tables S1a-b, S2 a-b**). The proportions of cases and controls exposed to >1 inhibitor were similar: 20/56 cases (36%) vs 12/30 controls (40%).

**Figure 2:**
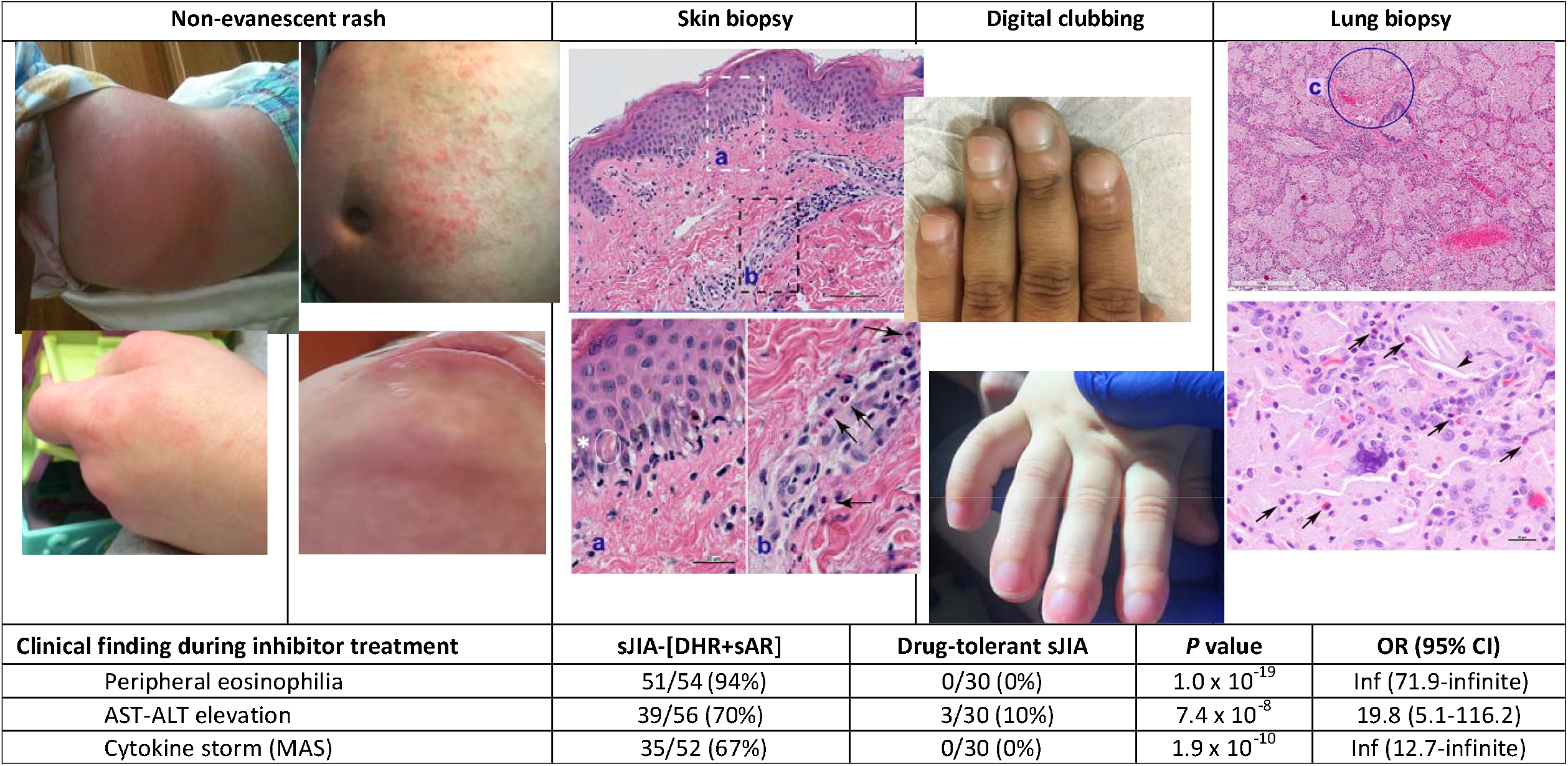
Unusual clinical features in inhibitor-treated sJIA patients. *P* value, Fisher exact probability test; OR (95%CI): odds ratio (95% confidence interval); sJIA, systemic juvenile idiopathic arthritis; DHR+sAR, Delayed hypersensitivity plus suspected anakinra reaction; MAS, macrophage activation syndrome, a form of secondary hemophagocytic lymphohistiocytosis; inf, infinite; AST-ALT elevation, aspartate aminotransferase (AST/SGOT) or alanine aminotransferase (ALT/SGPT) measuring >2x the upper limit of normal more than once, without alternative (non-drug) explanation. Two cases lacked information regarding eosinophilia. Cytokine storm during treatment was not evaluable in four cases, treated briefly (see methods). Images of non-evanescent rash, typically pruritic, are shown. Upper left: On rilonacept, erythema and prominent edema affecting knee; upper right: on tocilizumab, excoriated and areas of hyperpigmentation on abdomen; lower left: On canakinumab, erythematous, edematous rash on hand [similar rash on face and ear (not shown)]; lower right: On anakinra, erythema, edema and non-herpetic vesiculation on face. Skin biopsy of drug-associated rash shows vacuolar interface dermatitis and eosinophils. Higher power images (sections a, b) show lymphocytes, vacuolation at the dermal-epidermal junction, focal dyskeratotic keratinocytes (asterisk) and perivascular eosinophils (arrows). In DHR patients with diffuse lung disease, acute digital clubbing, which was often erythematous, frequently was the first indication of lung involvement, on anakinra (top) and on canakinumab (bottom). Lung biopsy significant for variant pulmonary alveolar proteinosis/endogenous lipoid pneumonia and arterial wall thickening (c). Higher power image (below) shows cholesterol clefts (arrowhead) and scattered eosinophils (arrows). 8/16 reviewed cases showed eosinophils in many fields (see supplementary methods). Increased lung eosinophils are consistent with DHR and also seen in various inflammatory diseases. Peripheral eosinophilia: Median (IQR) peak absolute eosinophil count in sJIA-[DHR+sAR] subjects was 2014/ul (1044,3593); peak eosinophil % of WBC was 20% (13,36). Three cases without eosinophilia were treated with uninterrupted high dose steroids (> 0.5mg/k/d). Details in Methods and additional clinical information in Tables S2a,b.

DHR features appeared during treatment at FDA-approved doses for autoinflammatory diseases. Clinical DHR features were distinguishable from an sJIA/AOSD flare and notably included eosinophilia and non-sJIA/AOSD rash (**figure 2**). Peripheral blood eosinophilia without other cause (e.g., pre-existing atopy) was observed in 51/54 (94%). In >50% of cases, eosinophilia was pronounced despite concurrent steroids. Non-evanescent drug eruptions were observed in 52/56 (93%). In 32/39 (82%) providing detail, rash included facial rash and/or edema, which are typical of DRESS.^2^ Skin biopsy reports (11 cases) described features of drug reaction, including interface dermatitis, dyskeratosis and eosinophilia. Although vacuolar interface dermatitis can be seen in sJIA^16^, the predominance of eosinophils favored interface drug eruption.^7^ In 39/56 (70%) sJIA/AOSD DHR cases, AST-ALT elevation was noted in the absence of macrophage activation syndrome (MAS) or other explanation. Also, cytokine storm during inhibitor treatment, which can be a manifestation of DRESS^1,2,18,19^, was significantly more common in DHR patients than in drug-tolerant sJIA/AOSD (p=8.5×10^−10^). When MAS occurred during drug treatment, eosinophilia typically preceded this by months, consistent with evolution of DRESS-associated features.^2,20^ Additional clinical details, including selected information about DLD subjects, are in **figure 2** and **tables S1, S2**.

DHR was often unrecognized, as reflected by continuation of inhibitor therapy after DHR or DRESS criteria were met. Only 17/56 (30%) of sJIA/AOSD patients with DHR discontinued IL-1/IL-6 inhibitors for ≥3 months without re-introduction. In this group, rash, eosinophilia and AST-ALT elevation resolved in all cases, consistent with the DHR diagnosis. In addition, their systemic and arthritic sJIA became easier to manage. For example, 10/17 (59%) discontinued steroids and only 2/17 cases required steroids >6 months after drug stop [median follow-up 14 months (IQR: 6,36)]. By contrast, of 31 cases known to continue inhibitors after initial DHR features, 8 died and only 17% of survivors were off steroids, despite longer median follow-up of 27 months (IQR: 16,53). Overall, we observed DHR/DRESS to IL-1 and IL-6 inhibitors across the spectrum of sJIA-like, sJIA and AOSD, in patients with or without complicating DLD, and stopping inhibitors led to improvement.

### Common HLA-DRB1*15 alleles are risk factors for DHR to IL-1 and IL-6 inhibitors

To test for an association with inhibitor-triggered DHR, we collected HLA data. Visual inspection of the data from sJIA-DHR subjects, both with and without DLD (**table S1a**), revealed a predominance of a common haplotype, HLA-DRB5*01:01∼DRB1*15:01∼DQA1*01:02∼DQB1* 06:02; no predominant HLA class I allele was observed (not shown). Using HLA-DRB1*15:01 as a haplotype proxy, we found an astonishingly high frequency (83%) of this allele among 29 self-identified White sJIA subjects with DHR vs 0% in 19 drug-tolerant White sJIA controls (p=7×10^−9^; OR lower bound=14.5; **tables 1, S1a-b**). Comparisons of the sJIA-DHR subjects [29 White and the subset of 14 genetically-matched, European (**figure S1**)] to the INCHARGE sJIA subjects of European descent (n=550) revealed a similarly striking association of HLA-DRB1*15:01 in sJIA-DHR (p=1×10^−10^and p=1×10^−7^, respectively; **table 1, table S4**).

Another 26 subjects with sJIA/DHR, 1 with sJIA/sAR, and 11 who were drug-tolerant had self-identified race/ancestry other than White. Although the sample was insufficient to perform within-group analyses, we noted a similarly striking pattern of HLA association. HLA-DRB1*15:01 was observed in 55% non-White subjects with DHR and 0% of drug-tolerant controls (**tables 1, S1a-b**). Other alleles of the DRB1*15 family are more often present in non-European populations, and these appear to be associated with DHR as well. Together, HLA-DRB1*15 alleles (specifically HLA-DRB1*15:01, *15:03, *15:06) were noted in 74% non-White subjects with sJIA/DHR compared to 18% drug tolerant controls (**table 1**). Comparison of all sJIA/DHR subjects (regardless of ancestry) to all sJIA subjects in the INCHARGE sJIA cohort (n=773) similarly showed 79% with HLA-DRB1*15 alleles in the sJIA/DHR group versus 24% in the sJIA INCHARGE cohort (**table 1**).

The White sJIA-DHR cohort also showed an expected HLA association with sJIA. The sJIA-associated HLA-DRB1*11:01 allele^21^ was found in sJIA-DHR European cases at a similar frequency to that of the INCHARGE sJIA cases, whereas it was enriched compared to INCHARGE controls (29% vs 10%; p=0.04; **table 2, table S4**). Notably, HLA-DRB1*15:01 was not associated with sJIA in the European INCHARGE cohort (**table 2**). Considering all ancestries, frequencies of HLA-DRB1*11:01 and HLA-DRB1*15:XX in sJIA-DHR and INCHARGE sJIA were consistent with the specificity of HLA-DRB1*11 association for sJIA and HLA-DRB1*15 association for DHR (**tables 1, 2, table S4**).

### Common HLA-DRB1*15 alleles are also likely risk factors for suspected anakinra reaction in KD

To determine whether HLA-linked DHR required sJIA-specific immune dysfunction, we studied a small cohort (n=19) of children with KD in a trial of 2-6 weeks of anakinra treatment.^12^ Four had suspected delayed anakinra reaction (sAR; **table S3**). We observed the same striking effect; 3/4 children with sAR carried HLA-DRB1*15 alleles (HLA-DRB1*15:01 and *15:03), whereas a different HLA-DRB1*15 allele, HLA-DRB1*15:02, was observed in 2/15 apparently drug-tolerant children (**tables 1, S1a-b**). Notably, HLA-DRB1*15:01 was not observed in any drug-tolerant KD subject (**table S1b**).

Finally, while the HLA-DRB1*15:01∼DQA1*01:02∼DQB1*06:02 haplotype is in near-complete linkage disequilibrium (LD) in European populations, analysis across ancestries, in which patterns of LD differ, can help to pinpoint the associated locus. Considering the entire sJIA-DHR+KD-sAR group, HLA-DRB1*15:01 was observed in 68% of subjects with DHR and was completely absent in drug-tolerant controls (**table 1**). In contrast, HLA-DQB1*06:02 was observed in 7% of controls, in the context of different haplotypes (**table 1, tables S1b**), suggesting HLA-DRB1 as the operative locus. It is important to note, however, that HLA-DRB5*01:01, an allele of a secondary HLA-DRB locus, is found on nearly all haplotypes with HLA-DRB1*15. We did not assay this locus in every subject, and we are not able to rule it out as the primary effector.

## Discussion

We have uncovered strong evidence for DHR/sAR to anakinra, canakinumab, rilonacept (inhibitors of IL-1) and to tocilizumab, an inhibitor of IL-6. The clinical picture of DHR is similar across these drugs and across DHR patients along the spectrum of sJIA-like disease, sJIA and AOSD. These patients meet classification criteria for DRESS, a potentially fatal, eosinophilic systemic syndrome that can lead to organ failure and can stimulate a cytokine storm.^1,2,18,19^ In relatively short-term exposure of KD patients to anakinra, a subset of patients also developed clinical manifestations consistent with drug reaction. In sJIA/AOSD and KD, the drug reactions are delayed type and differ from the immediate, anaphylactic reactions to tocilizumab we observed in association with DLD in sJIA.^10^ Although some subjects experienced both types of drug reactions, most did not, and we have not identified an inherited risk factor for anaphylaxis to tocilizumab.

Importantly, we also discovered a genetic risk factor for DHR to these inhibitors. We observed a very strong association of the HLA-DRB1*15:01 allele (and the linked HLA-DRB5*01:01 where measured) to DHR in White sJIA subjects; the numbers of cases, drug-tolerant controls and sJIA controls (INCHARGE) allowed rigorous analysis of this ancestry group. Our sample size was sufficient to detect what appears to be complete penetrance of the risk allele, as evidenced by its complete absence in drug-tolerant controls and the highly significant p-values we report. Although we were limited by the relative scarcity of non-White subjects in our sample, our findings also suggest that, in addition to HLA-DRB1*15:01, other alleles of the HLA-DRB1*15 family are linked to risk for DHR in these populations. The distribution of subjects with HLA-DRB1*15 argues the DHR risk applies across ancestry groups (**figure 3**), as found in some other HLA/DHR associations.^6^ Carriers of DRB1*15:01, *15:03, *15:06 alleles are common [27% (White), 15% (Hispanic), 27% (Black) and 16% (Asians) in US populations].^22^ Our current cohort does not allow analysis of HLA-DRB1*15:02, a high frequency allele in Asian populations. Approximately 20% of the DHR subjects do not carry the risk alleles; determining if other genetic factors confer risk will be important.

**Figure 3:**
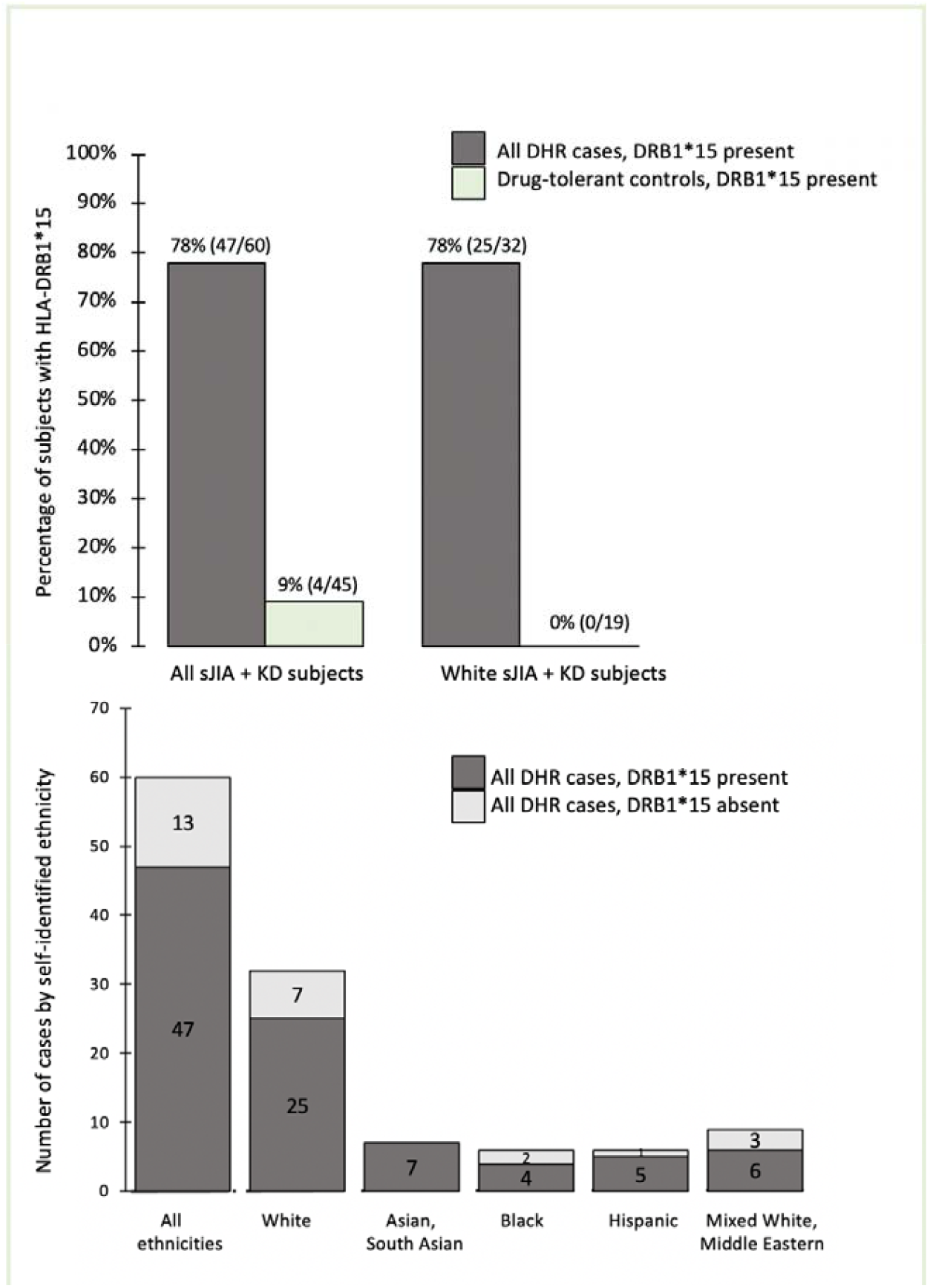
HLA-DRB1*15 is enriched in DHR across ethnicities. DHR, delayed hypersensitivity reaction, with or without diffuse lung disease, including suspected anakinra reaction; All DHR cases, systemic onset juvenile arthritis and Kawasaki disease; HLA-DRB1*15, any DRB1*15 allele present in cases or controls (*15:01, *15:02, *15:03, *15:06) Percentage of DHR cases and controls with HLA-DRB1*15 across ethnicities (upper, left) and in self-identified White cohort (upper, right). Lower panel shows numbers of cases with and without HLA-DRB1*15 in all ethnicities and in each self-identified ethnic group. In cases with DRB*3/4/5 allele information, all subjects with DRB1*15 (n=33) also carry DRB5*01:01. (table S1a)

Unusual aspects of this DHR/HLA association are its restriction to HLA class II^5,6^ and that the risk spans several inhibitors with different chemical structures (**figure S2**). The latter raises the possibility that an excipient common to these drugs and/or a molecule increased by inhibition of the intersecting IL-1 and IL-6 pathways creates a stimulatory HLA class II molecule, which activates CD4+ T cells. Several molecular mechanisms for the modification of HLA into an immunogenic moiety have been identified or proposed.^8^ A detailed picture of the pathogenesis of clinical DHR remains to be elucidated and may involve a complex interplay between viruses, HLA proteins, T cells, cytokine secretion and other genetic polymorphisms.^1,6,23^

The conditions for which these inhibitors may be used is a large and expanding group, including monogenic autoinflammatory diseases, rheumatoid arthritis (RA), polyarthritis, type 2 diabetes, gout, multisystem inflammatory syndrome in children, and cytokine storms associated with CAR-T therapy and COVID-19.^24-26^ These drugs are also in trials for multiple sclerosis, asthma, pericarditis, ST-segment–elevation acute myocardial infarction, heart failure with reduced ejection fraction and lung cancer.^25,26^ We found scattered reports of DRESS or hyper-eosinophilia with rash implicating these drugs in RA, polyarthritis, undifferentiated autoinflammatory disorder, giant cell arteritis and COVID-related cytokine storm (**table S5**). HLA typing was not included in these reports and will be important in future investigations.

Previous studies of IL-1 or IL-6 inhibitors do not mention possible DHR. However, it is possible that DHR was unrecognized. In a recent study of anakinra as first-line therapy for sJIA, 17% of subjects required high dose steroids for clinical deterioration or MAS.^27^ The pivotal trial of canakinumab for sJIA had a 19% non-response rate.^28^ A study of tocilizumab in RA had a 15% withdrawal rate for adverse events and/or failure to respond.^29^ In 24 COVID-19 patients treated with tocilizumab, post-treatment elevation of IL-6 levels identified the 25% who died.^30^ Further work is needed to determine if hypersensitivity contributes to the rates of drug failures.

There are several limitations to our study. First, our drug-tolerant control group was small. We addressed this limitation by using the INCHARGE sJIA cohort as a comparator, although the drug tolerance status of these subjects is unknown. Notably, however, unidentified sJIA/DHR cases among these subjects would mean the high odds ratio we observe is an underestimate of the true effect size. Second, our convenience cohort does not provide information on the prevalence of DHR with DLD as compared to DHR without DLD; indeed, the latter may be more common. Other limitations include those inherent to a retrospective observational design, such as missing data. For example, we lacked information to determine whether the DHR subset had a higher frequency of herpes virus reactivation, particularly HHV-6, as reported in DRESS.^2,6^ Lastly, and importantly, our convenience sample had under-representation of non-White subjects, limiting our genetic/HLA analyses. Validation and confirmation of our findings across ethnicities is an important next step.

An unanswered question is whether/how immune-mediated DHR links to the development of DLD in sJIA.^2,3^ The correlation between increasing use of IL-1 and IL-6 inhibitors and increasing recognition of DLD in sJIA argues for a relationship. Drug-induced interstitial lung disease, including PAP, are reported.^10,31-34^ It will be critical to determine if DLD in sJIA improves by withdrawing DHR-associated drugs, if there is a window of opportunity for this intervention, and if drug tolerance develops over time, especially with concurrent immune suppression. In light of our results, it seems unlikely that DLD is HLA-DRB1*15-associated, independent of DHR.

Given the high frequency of the risk alleles across populations, our findings encourage pre-prescription risk analysis, using readily available HLA testing. The HLA association is at least equivalent in effect to the association of HLA-B*57:01 with hypersensitivity to abacavir^35^, for which HLA screening is recommended. For individuals without the identified risk alleles, attention to signs of hypersensitivity is prudent. Similar to recent reports^36,37^, we observed onset of severe DHR as early as three days after first exposure but also after months of treatment. Thus, our results are relevant for short-term use of the implicated inhibitors and also highlight the need for continued surveillance for DHR over time. Further research is needed to determine underlying mechanisms for DHR to these inhibitors and its relevance in other conditions, particularly inflammatory diseases, in which DRESS/DHR is underrecognized.^2,18^

## Data Availability

All data referred to in the manuscript will be available upon reasonable request.

