## Supplementary Appendix for "IL-1 and IL-6 inhibitor hypersensitivity link to common HLA-DRB1*15 alleles"

This appendix has been provided by the authors to give readers additional information about their work.

**Supplementary Appendix**

**Table of Contents**

**Investigators**	3

**Acknowledgements**	4

**Supplementary Methods**	5

Approvals	5

Subjects and clinical data collection	5

Ancestry data collection	6

Lung histopathology	6

DNA extraction methods	6

HLA genotyping	7

*HLA genotyping via next-generation sequencing (NGS)*

*HLA genotyping via reverse sequence-specific oligonucleotide (rSSO)*

*HLA genotyping from WES data*

Data analyses	8

**Supplementary Tables**	9

Notes regarding the tables	9

Table S1a: HLA-DQ and -DR in cases with delayed hypersensitivity reaction (DHR)	10

Table S1b: HLA-DQ and -DR in drug-tolerant controls	12

Table S2a: Clinical features in sJIA cases with DHR	13

Table S2b: Clinical features in sJIA drug-tolerant controls	15

Table S3: Clinical features in Kawasaki disease cases and controls	16

Table S4: Association analysis for HLA alleles in sJIA-DHR subjects vs. INCHARGE collection	17

Table S5: Published case reports of delayed drug reactions implicating IL-1 or IL-6 inhibitors	17

**Supplementary Figures**	18

Figure S1: Ancestral matching of INCHARGE sJIA GWAS cases and controls with sJIA-DHR subjects	18

Figure S2: Chemical structures of cytokine blockers implicated in HLA-DRB1*15-associated DHR	19

**References for Supplementary Information**	20

**Investigators**

Jill A. Hollenbach (University of California San Francisco, San Francisco, CA, USA)

Michael J. Ombrello (National Institute of Arthritis and Musculoskeletal and Skin Diseases, Bethesda, MD, USA)

Adriana H. Tremoulet (University of California San Diego, San Diego, CA, USA)

Gonzalo Montero-Martin, MsC (Stanford University, Stanford, CA, USA)

Sampath Prahalad (Emory University School of Medicine, Children’s Healthcare of Atlanta, Atlanta, GA, USA)

Scott Canna (University of Pittsburgh, Pittsburgh, PA, USA)

Chisato Shimizu (University of California San Diego, San Diego, CA, USA)

Gail Deutsch (University of Washington, Seattle, WA, USA)

Serena Tan (Stanford University, Stanford, CA, USA)

Elaine F. Remmers (National Human Genome Research Institute, Bethesda, MD, USA)

Dimitri Monos (University of Pennsylvania, Philadelphia, PA, USA)

Omkar K. Phadke (Emory University School of Medicine, Children’s Healthcare of Atlanta, Atlanta, GA, USA)

Jianpeng Xu (Stanford University, Stanford, CA, USA)

Jaime S. Rosa Duque (The University of Hong Kong, Hong Kong Special Administrative Region, China)

Gilbert T. Chua (The University of Hong Kong, Hong Kong Special Administrative Region, China)

Vamsee Mallajosyula (Stanford University, Stanford, CA, USA)

Debopam Ghosh (Stanford University, Stanford, CA, USA)

Ann Marie Szymanski (National Institute of Arthritis and Musculoskeletal and Skin Diseases, Bethesda, MD, USA)

Danielle Rubin (National Institute of Arthritis and Musculoskeletal and Skin Diseases, Bethesda, MD, USA)

Jane C. Burns (University of California San Diego, San Diego, CA, USA)

Marcelo A. Fernandez-Vina (Stanford University, Stanford, CA, USA)

Elizabeth D. Mellins (Stanford University, Stanford, CA, USA)

Vivian E. Saper (Stanford University, Stanford, CA, USA)

**Drug hypersensitivity consortium**

Rabheh Abdul Aziz (State University of New York, Buffalo, New York, USA)

Roberta Berard (Children’s Hospital, London Health Sciences Centre, London, Ontario, Canada)

Catherine A. Bingham (Pennsylvania State University College of Medicine, Hershey, Pennsylvania, USA)

Alexis D. Bonaparth (Columbia University Medical Center, New York, New York, USA)

Alicia Casey (Boston Children's Hospital, Boston, Massachusetts, USA)

Elaine Cassidy (University of Pittsburgh Medical Center, Pittsburgh, Pennsylvania, USA)

Kathleen P. Collins (Stanford University, Stanford, California, USA)

Ian Ferguson (Yale University Medical School, New Haven, Connecticut, USA)

Steven I. Goodman (Arthritis Associates of South Florida, Delray Beach, Florida, USA)

Alexei A. Grom (University of Cincinnati College of Medicine, Cincinnati, Ohio, USA)

Melissa Hazen (Boston Children's Hospital, Boston, Massachusetts, USA)

Timothy Hahn (Pennsylvania State University College of Medicine, Hershey, Pennsylvania, USA)

Alice Hoftman (University of California Los Angeles Medical Center, Los Angeles, California, USA)

Maria Ibarra (School of Medicine, University of Missouri Kansas City, Kansas City, Missouri, USA)

Rita Jerath (Augusta University, Augusta, Georgia, USA)

Daniel J. Kingsbury (Randall Children’s Hospital at Legacy Emanuel, Portland, Oregon, USA)

Marisa S. Klein-Gitelman (Northwestern University Feinberg School of Medicine, Chicago, Illinois, USA)

Khanh Lai (University of Utah Health Sciences Center, Salt Lake City, Utah, USA)

Sivia Lapidus (Hackensack University Medical Center, Hackensack, New Jersey, USA)

Roberto Mendoza-Londono (Dept. of Paediatrics, The Hospital for Sick Children, University of Toronto, Canada)

Karen Onel (Hospital for Special Surgery, New York, New York, USA)

Maria Perez (Cook Children’s Medical Center, Fort Worth, Texas, USA)

Suhas M. Radhakrishna (University of California San Diego, CA)

Adam Reinhardt (Boystown National Research Hospital, Boystown, Nebraska, USA)

Mona Riskalla (University of Minnesota, Minneapolis, Minnesota, USA)

Johannes Roth (Children’s Hospital of Eastern Ontario, Ottawa, Ontario, Canada)

Natalie Rosenwasser (Seattle Children's Hospital, University of Washington, Seattle, Washington, USA)

Nadine Saad (Hospital for Special Surgery, New York, New York, USA)

Grant S. Schulert (University of Cincinnati College of Medicine, Cincinnati, Ohio, USA)

Susan Shenoi (Seattle Children's Hospital, University of Washington, Seattle, Washington, USA)

Judith A. Smith (University of Wisconsin Madison School of Medicine & Public Health, Madison, Wisconsin, USA)

Jennifer Soep (University of Colorado School of Medicine, Denver, Colorado, USA)

Cory Stingl (Children’s Hospital of Philadelphia, Philadelphia, Pennsylvania, USA)

Matthew Stoll (University of Alabama, Birmingham, Alabama, USA)

Melissa Tesher (University of Chicago Medical Center, Chicago, Illinois, USA)

Lawrence Zemel (University of Connecticut School of Medicine, Hartford, Connecticut, USA)

**Acknowledgements**

This work utilized *in silico* genetic data generated by the International Childhood Arthritis Genetics Consortium; we gratefully acknowledge the INCHARGE investigators. We also thank Debra D. Hiraki, Farkhondeh Ziaei, Linda Gojenola and Drs. James Zehnder and Bing M. Zhang of Stanford Pathology Department for assisting with sample procurement, processing and HLA genotyping, Jamie L. Duke (Children’s Hospital of Philadelphia) for assistance with HLA sequence analysis, Elizabeth Moreno (Rady Children’s Hospital) for assistance with Kawasaki disease samples and clinical data, Lori Ponder (Children’s Healthcare of Atlanta) for patient consents and facilitating sample collection, and Carol Lake (National Institute of Arthritis and Musculoskeletal and Skin Diseases) for accessing and aggregating clinical and laboratory data. We thank the Stanford Blood Center for support performing HLA tests and the following individuals at Stanford University: Drs. Tzielan Lee, Joyce Hsu, Imelda Balboni, Rajdeep Puni, Uptej Khalsa, Claudia Macaubas and Bernice Kwong for facilitating this project and Dr. Ann Leung for reviewing CT scans.

**Supplementary Methods**

**Approvals**

This study was approved by the Institutional Review Board (IRB), Stanford University, (#13932 and #34679). Local IRB from contributing institutions were obtained as needed, based on local practice. Informed consent was obtained, as required, for HLA genotyping or sharing genetic data. Subjects evaluated at the National Institutes of Health (NIH) were enrolled in a study of the natural history, genetics and pathophysiology of systemic arthritis (NCT03510442) and were evaluated with family-based whole exome sequencing (WES). The study was approved by the IRB of the NIH, and parents and subjects provided written informed consent/assent, as appropriate. Subjects with Kawasaki disease (KD) were enrolled at Rady Children’s Hospital San Diego in a Phase I/IIa, dose-escalation trial of anakinra for intensification of initial therapy in patients with coronary artery aneurysms on the first echocardiogram (NCT 02179853). The study was approved by the IRB at the University of California San Diego, and parents and subjects signed written consent/assent as appropriate.

**Subjects and clinical data collection**

sJIA patients (n=76) met an operational case definition, developed by expert consensus as a modification of the ILAR (International League of Associations for Rheumatology) sJIA classification criteria.^1^ sJIA-like patients (n=7) were those managed clinically like sJIA, but not meeting modified ILAR criteria. AOSD patients (n=3) met Yamaguchi criteria.^2^ Onset age at sJIA diagnosis was evaluated by age group (<2.5yrs, 2.5-10, >10). Patients with onset age ≥ 18 yrs were classified as AOSD; 5 DHR cases and 2 sJIA controls had onset age ≥ 16 yrs. KD subjects (n=19) met inclusion criteria, for a Phase I/IIa trial of brief anakinra treatment;^3^ these criteria included American Heart Association criteria for KD^4^ and evidence coronary artery aneurysms. Anakinra doses ranged from 2-8 mg/kg/day. After 2 weeks of anakinra, if aneurysms were persistent, treatment continued for 4 weeks.^3^ In the 19 KD subjects studied, treatment duration ranged from 9 to 46 days. For some analyses, comparator populations were sJIA cases (without known lung disease) and healthy control subjects from the International Childhood Arthritis Genetics Consortium (INCHARGE) genome-wide association study.^5^ Their drug tolerance status was unknown.

Clinical information was collected using REDCap electronic capture tools^6^, hosted at Stanford University. Subjects included cases with delayed hypersensitivity reactions (DHR) implicating inhibitors of IL-1 or IL-6 with or without the development of a diffuse parenchymal lung disease (DLD). We previously reported on sJIA cases with parenchymal lung disease, which we called “LD”. In that study, we enrolled any sJIA subject with parenchymal lung disease.^7^ Here we have used a new term, DLD, which refers to a specific type of parenchymal lung disease that occurs in the context of DHR to IL-1/IL-6 inhibitors. When biopsied, lung tissue shows variant PAP/ELP (see lung histopathology below). Among the 105 subjects in the present study, 32 had sJIA, DLD and DHR to one or more of anakinra, canakinumab, rilonacept and tocilizumab. Six subjects had sJIA-like disease, DHR and DLD, and 1 had DLD, DHR, AOSD. Of 46 previously reported sJIA cases with LD after drug exposure^7^, all 20 with genetic information classified as DHR/DLD and were included in this analysis. Seventeen sJIA patients and 1 sJIA-like patient had DHR or sAR (n=1) to IL-1 or IL-6 inhibitors without DLD. All DHR cases occurred between 2006 and November 2020.

DRESS-related AST-ALT elevation was defined as aspartate aminotransferase (AST/SGOT) or alanine aminotransferase (ALT/SGPT) measuring >2x the upper limit of normal more than once, without macrophage activation syndrome (MAS), a cytokine storm syndrome known to complicate sJIA^8^, infection or other documented non-DHR causes. Skin biopsy reports were available for 11 cases [sJIA-DHR/DLD (6), sJIA-DHR (5)], and a representative skin biopsy was examined by a pathologist (ST). DLD diagnosis required biopsy or autopsy showing variant pulmonary alveolar proteinosis/endogenous lipoid pneumonia (vPAP/ELP) or chest CT with diffuse bilateral interstitial abnormalities.^7^ Histopathologic analysis by a pediatric pulmonary pathologist (GD) is described below. Peripheral eosinophilia during inhibitor treatment was defined as an absolute eosinophil count (AEC) ≥500. To qualify as MAS during inhibitor treatment, drug exposure >4 weeks was required (Table S2).

**Ancestry data collection**

Ancestry was determined by self-identification. A subset of subjects also underwent ancestry-matching analysis for comparison with INCHARGE subjects (see Data Analysis). Ancestry data were provided by case reporters based on parents’ response and then selection from a list of categories (White, Black, Hispanic, Asian, South Asian, Middle Eastern, noting mixed parentage).

| **Ancestry groups** | |
| --- | --- |
| **Group** | **Definition** |
| White | Self-identification as White or White European |
| Middle Eastern | Egypt, Oman, Yemen, Qatar, United Arab Emirates, Iran, Bahrain, Syria, Jordan, Turkey, Lebanon, Saudi Arabia, Kuwait, Iraq, Israel |
| Hispanic | Self-identification as Hispanic, Latinx, Central or South American |
| South Asian | Afghanistan, India, Pakistan, Bangladesh, Sri Lanka, Nepal, Bhutan, Maldives |
| Asian | Central, Northern, Eastern, Western and South Eastern Asia |
| Black | Self-identification as Black or Black-African |

**Lung histopathology**

Hematoxylin and eosin (H&E) slides of lung tissue from 16 sJIA-DHR subjects were reviewed (GD); 4 others had pathology reports from treating institutions with similar findings (table S2). All cases demonstrated vPAP/ELP, characterized by alveolar filling with eosinophilic proteinaceous material admixed with a variable degree of cholesterol clefts and foamy macrophages, as described. Some degree of inflammation (neutrophils, lymphocytes, plasma cells, eosinophils) was frequently present, as was wall thickening of pulmonary arteries, as described.^7^ Increased lung eosinophils are seen in DHR^9^ and various inflammatory diseases, including rheumatologic conditions.^10^ Grading of eosinophils on biopsy used a semi-quantitative grading scale to assess the number of eosinophils on H&E stain at X20 magnification; at least 10 fields assessed. 0=no eosinophils, 1=scattered eosinophils in few fields, 2=scattered eosinophils in many fields with rare aggregates, 3=scattered eosinophils in most fields with several aggregates, 4=numerous aggregates of eosinophils. 3 cases were graded 0; 5 cases at 1; 8 cases at 2-3; 0 cases at 4. All subjects with tissue graded 0 were on high dose (1-2mg/kg/day) steroids (not shown).

**DNA extraction methods**

Different DNA extraction methods were used, as follows. Genomic DNA (gDNA) was extracted from peripheral blood samples of subjects recruited at Stanford University using the QIAsymphony DNA Mini Kit (based on silica matrix solid-phase purification) automated protocol, per manufacturer's instructions (Qiagen, Hilden, Germany). For subjects recruited at the NIH, gDNA was extracted from peripheral blood samples using the Maxwell 16 DNA Purification Kits and the automated Maxwell 16 instrument, per manufacturer’s instructions (Promega, Madison, WI, United States). For the patients with KD, gDNA was extracted from peripheral blood samples using the Wizard Genomic DNA purification kit, per manufacturer's instructions (Promega, Madison, WI, USA). In the case of formalin-fixed paraffin-embedded (FFPE) samples, gDNA was extracted using the QIAamp DNA FFPE tissue kit (Qiagen, Hilden, Germany), following the standard protocol of Stanford Molecular Pathology. Quantity and quality of extracted DNA samples were measured by spectrophotometry using the NanoDrop 2000 instrument (Thermo Fisher Scientific, Waltham, MA, USA) or by fluorometric quantification of double-stranded DNA using a Qubit 3 instrument (Thermo-Fisher Scientific, Waltham, MA, USA).

**HLA genotyping**

*HLA genotyping via next-generation sequencing (NGS)*

DNA samples were typed for HLA class I (*HLA-A, HLA-B* and *HLA-C*) and class II (*HLA-DPA1, HLA-DPB1, HLA-DQA1, HLA-DQB1, HLA-DRB1, HLA-DRB3, HLA-DRB4,* and *HLA-DRB5*) loci using the MIA FORA NGS FLEX high-throughput semi-automated typing protocol (Immucor, Inc., Norcross, GA, USA) per manufacturer’s instructions, as initially reported in Wang, C. et al.^11^ This NGS method allowed full-length coverage for HLA class I genes and extensive coverage for class II genes amplified by long range PCR.^12^ After DNA library preparation steps, pooled sample libraries were sequenced at a final concentration of 1.3 pM spiked with 20pM PhiX on the Illumina MiniSeq instrument using 150 cycle paired-end kits (Illumina, Inc., San Diego, CA). NGS sequencing reads stored as fastq files were uploaded into the MIA FORA NGS FLEX v4.5 HLA genotyping software (Immucor, Norcross, GA) for: building phased consensus sequences; establishing their respective alignment to reference sequences (according to IPD-IMGT/HLA v3.36.0 database, https://www.ebi.ac.uk/ipd/imgt/hla/ and to MIA FORA FLEX internal reference database of cloned and in-silico HLA allele sequences, https://www.immucor.com/en-us/Products/Pages/MIA-FORA-NGS.aspx).

*HLA genotyping via reverse sequence-specific oligonucleotide (rSSO) method*

Some extracted DNA samples were tested for *HLA-DQA1*, *HLA-DQB1*, *HLA-DRB1*, *HLA-DRB3*, *HLA-DRB4*, and *HLA-DRB5* loci via reverse sequence-specific oligonucleotide (rSSO) DNA typing method using the LABType SSO HLA semi-automated typing protocol (One Lambda, Inc., Canoga Park, CA) per manufacturer’s instructions (https://www.onelambda.com/en/product/labtype-sso.html). HLA genotyping data obtained by rSSO was analyzed according to IPD-IMGT/HLA v3.37.0 release database (https://www.ebi.ac.uk/ipd/imgt/hla/).

*HLA genotyping from WES data*

36 subjects had whole exome sequencing (WES) data from which HLA genotype was extracted. WES data from 14 subjects were analyzed at Stanford using HLAreporter.^13^ In brief, a preliminary high-efficiency mapping of the reads was achieved using Burrows-Wheeler Aligner^14^ against a comprehensive reference panel generated from all known HLA alleles in the IMGT/HLA database, followed by a *de novo* assembly of gene-specific mapped reads to contigs using Targeted Assembly of Short Sequence Reads.^15^ Typically, contigs with an average coverage-depth of ≥5-fold were picked for HLA allele assignment. Subsequently, these contigs were matched stepwise against reference databases of HLA alleles, followed by HLA allele calling for HLA-A, -B, -C, -DRB1, -DRB3/4/5, -DQA1, -DQB1, -DPB1. WES data from 24 subjects were analyzed at the NIH using HLA:LA^16^, a highly accurate graph-based method for extracting HLA alleles from exome and low-coverage whole genome sequencing data. Briefly, input reads were linearly aligned to Population Reference Graph (PRG) reference haplotypes, which include eight GRCh38 MHC haplotypes and all IMGT exonic and genomic sequences, using BWA-MEM.^17^ Linear alignments were projected onto the PRG and the alignments were optimized in a stepwise process. Contigs overlapping the MHC were identified using nucmer^18^, and each contig was annotated according to the HLA haplotype with the highest alignment score. HLA sequences were extracted according to genomic position, and HLA typing was performed by minimum edit distance-based matching with the IMGT database. Three subjects were analyzed at Stanford and the NIH, with identical results. For subjects from the INCHARGE sJIA GWAS cohort, HLA alleles at the 8 classical HLA loci (*HLA-A, HLA-B, HLA-C, HLA-DPB1, HLA-DPA1, HLA-DRB1, HLA-DQA1, HLA-DQB1*) were determined by HLA imputation with SNP2HLA software, as previously described.^19^ WES data from 1 subject and both parents were analyzed at The Children’s Hospital of Philadelphia (DM) using Explore (Omixon, Budapest, Hungary). Briefly, the fastq files were filtered in Explore for reads deriving from 35 HLA genes/pseudogenes within the MHC (*HLA-A, HLA-B, HLA-C, HLA-DMA, HLA-DMB, HLA-DOA, HLA-DOB, HLA-DPA1, HLA-DPA2, HLA-DPB1, HLA-DPB2, HLA-DQA1, HLA-DQB1, HLA-DRA, HLA-DRB1, HLA-DRB2, HLA-DRB3, HLA-DRB4, HLA-DRB5, HLA-DRB6, HLA-DRB7, HLA-DRB8, HLA-DRB9, HLA-E, HLA-F, HLA-G, HLA-H, HLA-J, HLA-K, HLA-L, HLA-T, HLA-U, HLA-V, HLA-W, HLA-Y*). The filtered reads were analyzed using Explore, targeting the same panel of MHC genes using both an exon-only and full gene analysis modes. Genotype data were reported for classical HLA loci (*HLA-A, HLA-B, HLA-C, HLA-DPA1, HLA-DPB1, HLA-DQA1, HLA-DQB1, HLA-DRB1, HLA-DRB3, HLA-DRB4,* and *HLA-DRB5*). Discrepancies between the two methods were resolved manually. Haplotype analysis of the trio confirmed the genotype of the patient. All analysis was conducted using the IMGT/HLA database version 3.38.0.

**Data analysis**

Association testing of HLA alleles in sJIA-DHR compared to the INCHARGE datasets. Principal component analysis (PCA) was performed with SNP & Variation Suite 8 (SVS8; Golden Helix, Bozeman, MT, USA), as previously described.^19^ Association testing with the INCHARGE collection was performed with SVS8, using logistic regression under the dominant model and including sex and ancestral information as covariates (**table S4**).

**Supplementary Tables**

**Notes regarding the tables:**

In tables S1, S2, S3, S4, sections with data on *cases* are shaded in blue

and those with data on *controls* are shaded in green.

For Supplementary Tables S1a and S2a, footnotes are provided on the page following the table.

All references are provided at the end of the Supplementary Appendix.

**Table S1a: HLA-DQ and -DR in cases with delayed hypersensitivity reaction (DHR)***

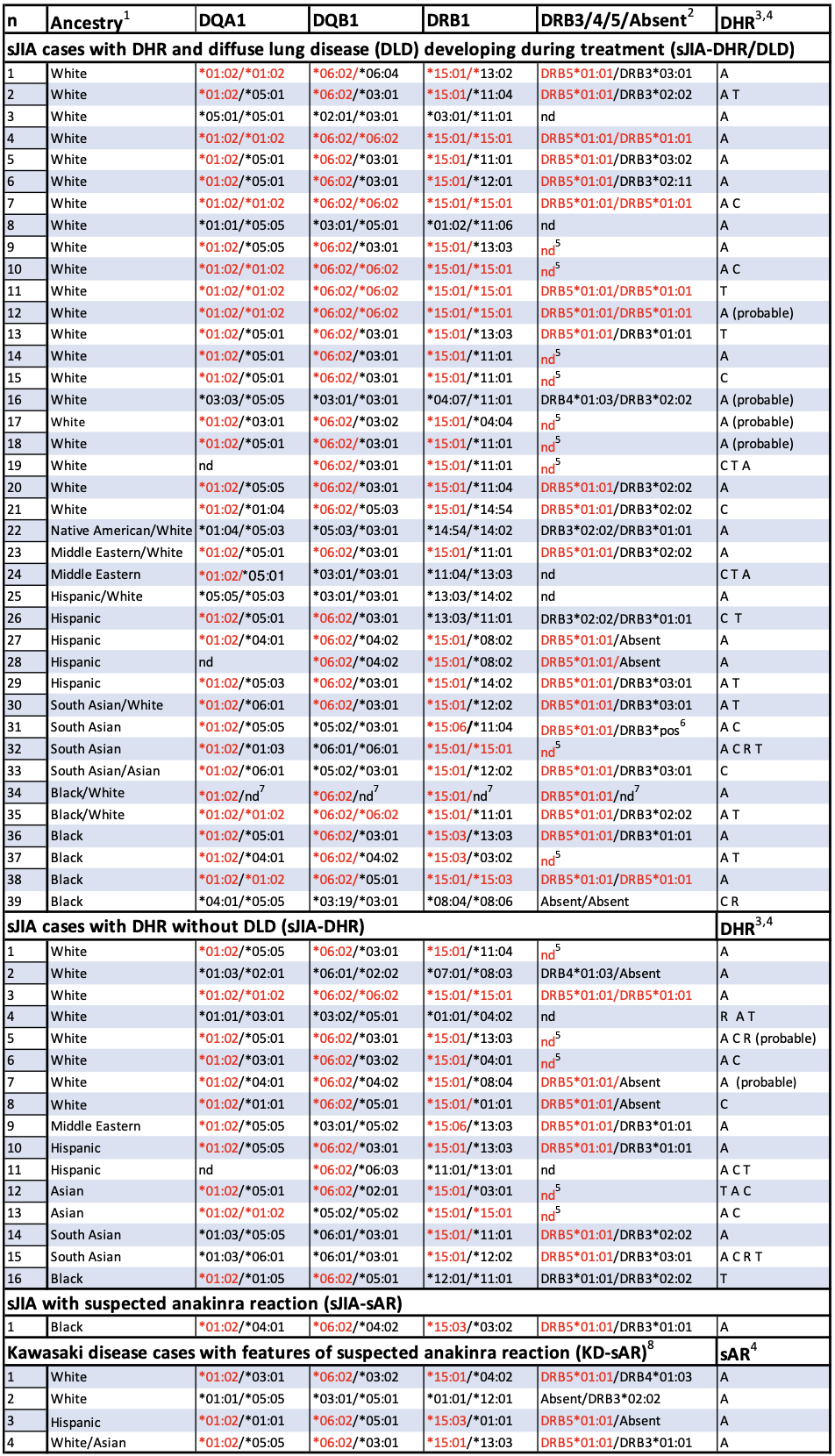

* An independent HLA class I association was not found (data not shown). Red indicates DHR/sAR risk alleles.

nd, not determined; DHR, delayed hypersensitivity reaction; sAR, suspected (delayed type) anakinra reaction.

**Footnotes for Table S1a**

^1^Ancestry self-identification as White, Hispanic, South Asian, Asian or Black, with mixed parentage noted.

^2^Absent: Absence of HLA-DRB3/4/5 when HLA-DRB1*01/*08/*10 are present^20^

^3^DHR by clinical diagnosis and/or 'definite' by RegiSCAR^21^; (probable): 'probable' by RegiSCAR, each with continuous high dose steroids, 1 with drug-induced liver injury and limited other data.

^4^Implicated drug for DHR: A, anakinra	C, canakinumab	R, rilonacept, T, tocilizumab

^5^Per http://17ihiw.org/17th-ihiw-ngs-hla-data/ and references.^22,23^ DRB5*01:01 is tightly linked to DRB1*15:01, 15:03, *15:06.

^6^Does not genotype due to crossmapping.

^7^Imputed as one DRB1*15:01 haplotype by single nucleotide polymorphism (SNP)^24^

^8^ Clinical information is provided on table S4.

**Table S1b: HLA-DQ and -DR in drug-tolerant controls**

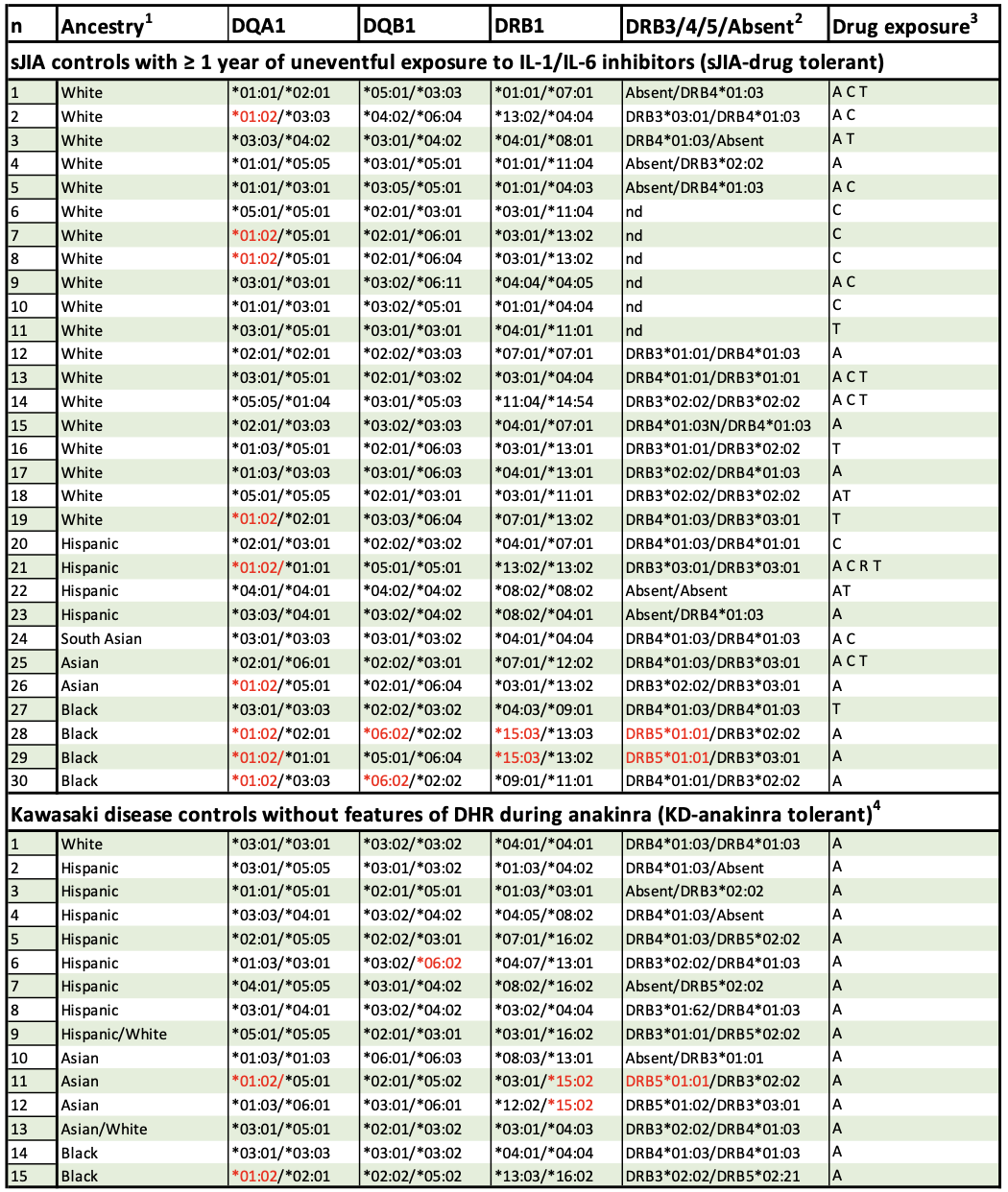

**S1a** HLA class I loci were typed in the majority of these subjects (not shown). Red indicates DHR/sAR risk alleles.

^1^Ancestry self-identification as White, Hispanic, South Asian, Asian or Black, with mixed parentage noted.

^2^Absent: Absence of HLA-DRB3/4/5 when HLA-DRB1*01/*08/*10 are present.^20^ ^Degenhardt^

^3^A, anakinra	C, canakinumab	R, rilonacept, T, tocilizumab

^4^ Clinical information is provided on table S4.

**Table S2a: Clinical features in sJIA cases with DHR**^1^
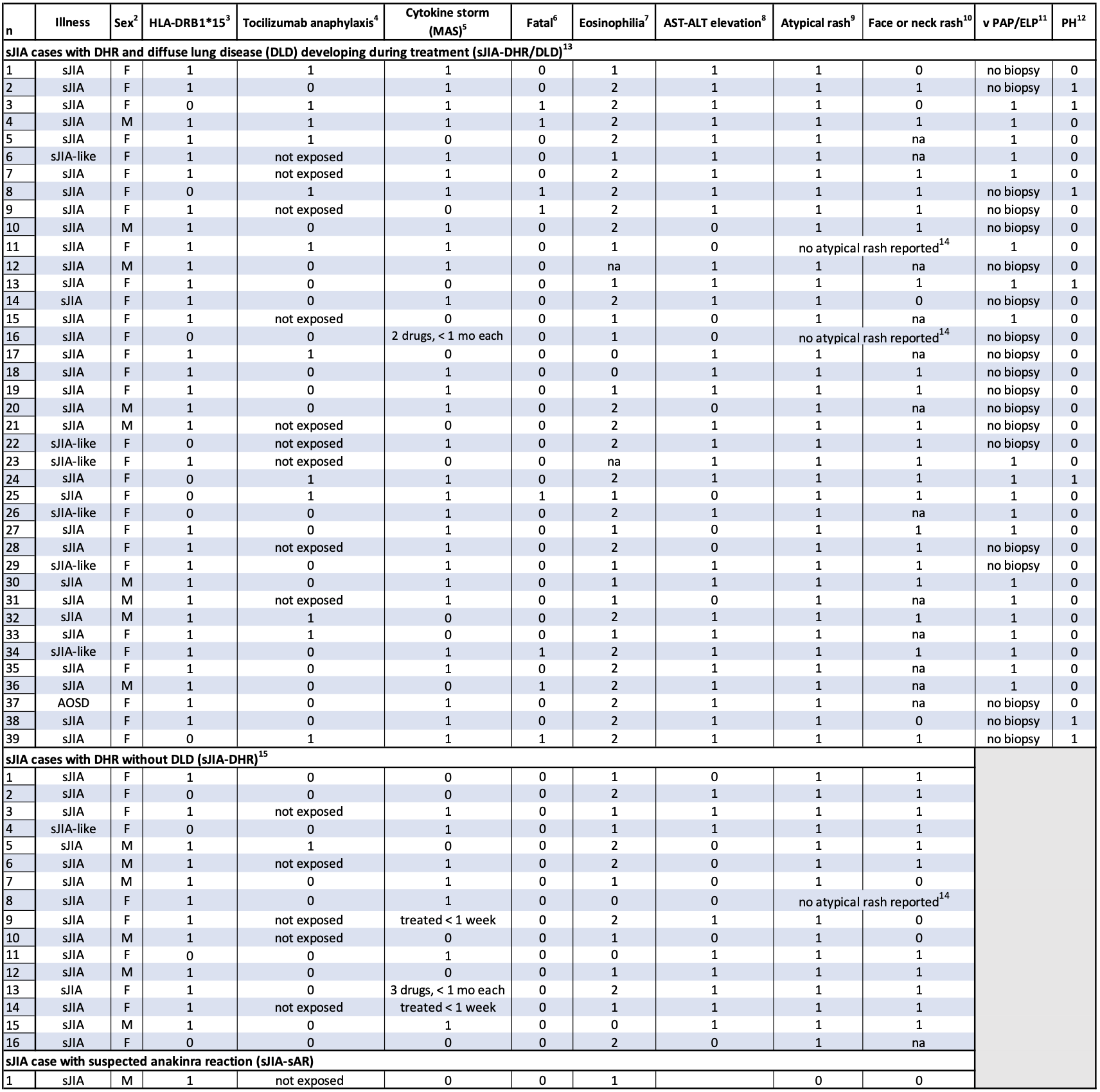

na, not available; 0=absent 1=present, except for eosinophilia, which is coded as per footnote 6; MAS, macrophage activation syndrome; PH, pulmonary hypertension; sJIA, systemic onset juvenile arthritis; sJIA-like, does not fulfill sJIA classification criteria and treated as sJIA; AOSD, adult onset Still's disease

**Footnotes for Table S2a**

^1^Clinical features during treatment with the drug implicated in DHR.

^2^No significant difference in sex frequency in cases (sJIA-DHR /DLD+sJIA-DHR) and controls (sJIA drug-tolerant): F vs M: OR 1.8 (95% CI 0.7,4.6), p=0.22.

^3^Presence of HLA-DRB1*15 (see table S1a).

^4^Immediate anaphylactic reaction (not DHR) during tocilizumab administration; all subjects discontinued drug. This reaction was not associated with HLA-DR 15: 8/30 (27%) HLA-DRB1*15 vs 5/11 (45%) without HLA-DRB1*15, p=0.14 OR 0.3 (0.07,1.27).

^5^MAS during inhibitor treatment occurred in cases continuing drug after initial DHR features. Omitted are 4 cases with very brief exposure, as indicated (range 4 days-4 weeks); Drug was stopped after DHR features began, none had MAS.

^6^Fatalities (7/7) occurred during continuation or restart of implicated drug. Time from DLD to data close ≥7 months.

^7^1= peak absolute eosinophil count (AEC) > normal limit <1500/ul (eosinophilic); 2= AEC ≥ 1500/ul (hyper-eosinophilic).^25^ For sJIA-DHR/DLD and sJIA-DHR, respectively, median (IQR) peak AEC = 2110/ul (1205,3945) and 1345/ul (930, 2500) and % eosinophil =24% (14,43) and 13% (11,23), despite concurrent steroids in >60% of subjects. sJIA-sAR peak AEC 1042 and 15.5% peak eosinophils. Cases without eosinophilia had uninterrupted steroid treatment.

^8^In the absence of MAS, elevated aspartate aminotransferase (AST) or alanine aminotransferase (ALT) > 2x the upper limit of normal on > 1 occasion during inhibitor treatment.

^9^Non-evanescent rash, often intensely pruritic, variably including hyperpigmentation, eczema, angioedema, confluent erythema, serpiginous, vesicular and flagellate dermatologic patterns. Skin biopsy reports, provided for 11 cases, were all consistent with drug reaction (interface dermatitis, dyskeratosis, eosinophilia (see fig. 2).

^10^Rash involving the face or neck is highly prevalent (in ~80%) in DRESS-type drug reactions.^26,27^

^11^Variant pulmonary alveolar proteinosis/endogenous lipoid pneumonia with notable presence of lymphocytoplasmic infiltrate and vascular changes.^2^

^12^Pulmonary hypertension by echocardiogram or cardiac catheterization.

^13^Time from first feature of DHR until recognition of DLD: 1 mo-2.3 yrs.

^14^Subjects without atypical rash had uninterrupted high dose steroid treatment.

^15^Median follow-up from first DHR feature to data close was 3.9 yrs (range 0.2, 8.7).

Shomali^25^ Saper ^7^

**Table S2b: Clinical features in sJIA drug-tolerant controls^1^**

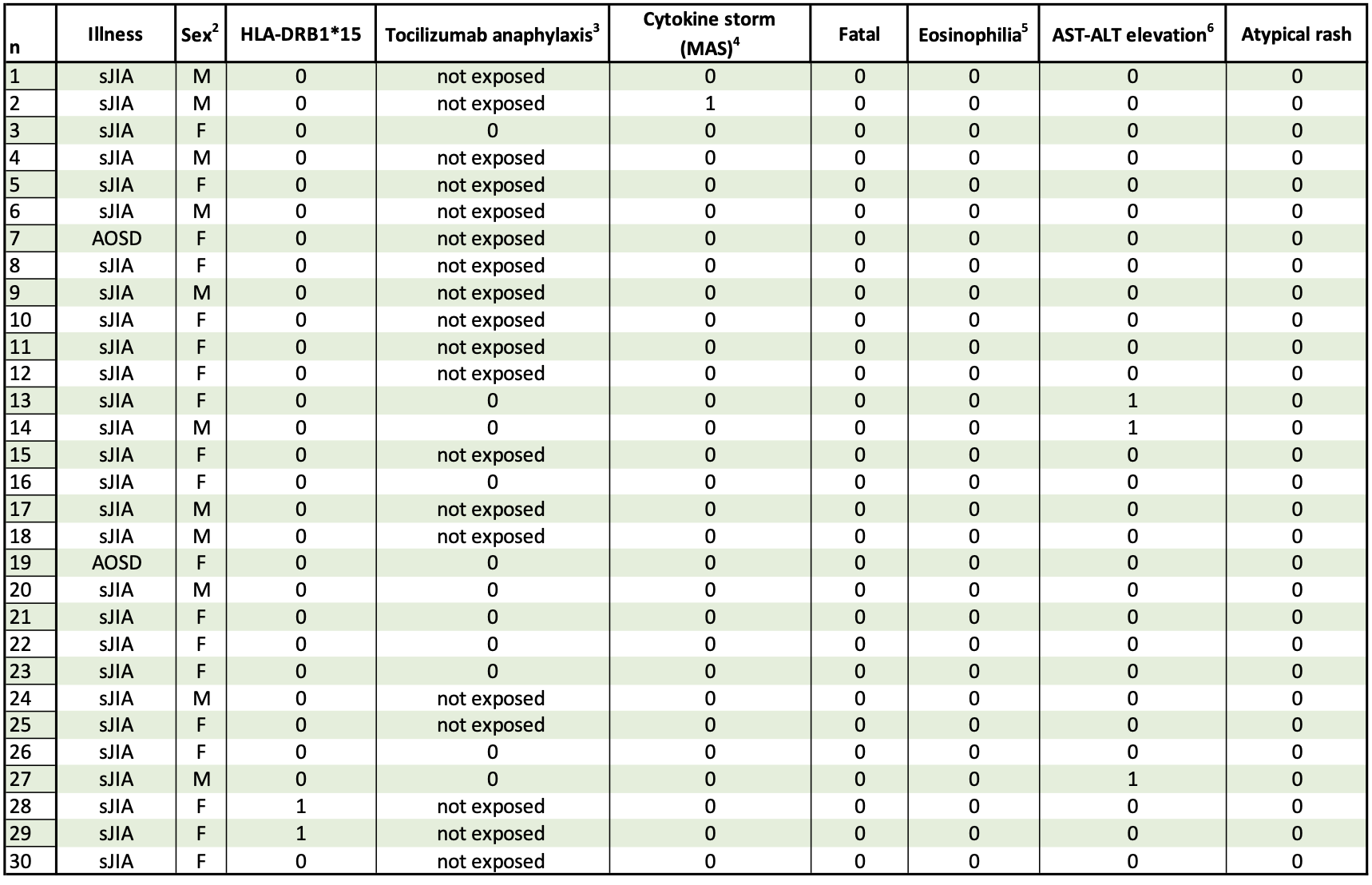

0=absent 1=present; sJIA, systemic onset juvenile arthritis; AOSD, adult onset Still's disease; MAS, macrophage activation syndrome

^1^Treated uneventfully for ≥1 yr. None with non-evanescent atypical rash.

^2^No significant difference in sex frequency in cases (sJIA-DHR /DLD+sJIA-DHR) and drug-tolerant controls (table S2).

^3^Immediate anaphylactic reaction during tocilizumab administration.

^4^Cytokine storm (macrophage activation syndrome) during treatment with inhibitors.

^5^Peak absolute eosinophil count (AEC) > normal limit occurring during treatment with inhibitors.

^6^In the absence of MAS, elevated aspartate aminotransferase (AST) or alanine aminotransferase (ALT) > 2x the upper limit of normal on > 1 occasion during inhibitor treatment.

**Table S3: Clinical features in Kawasaki disease cases and controls**^1^

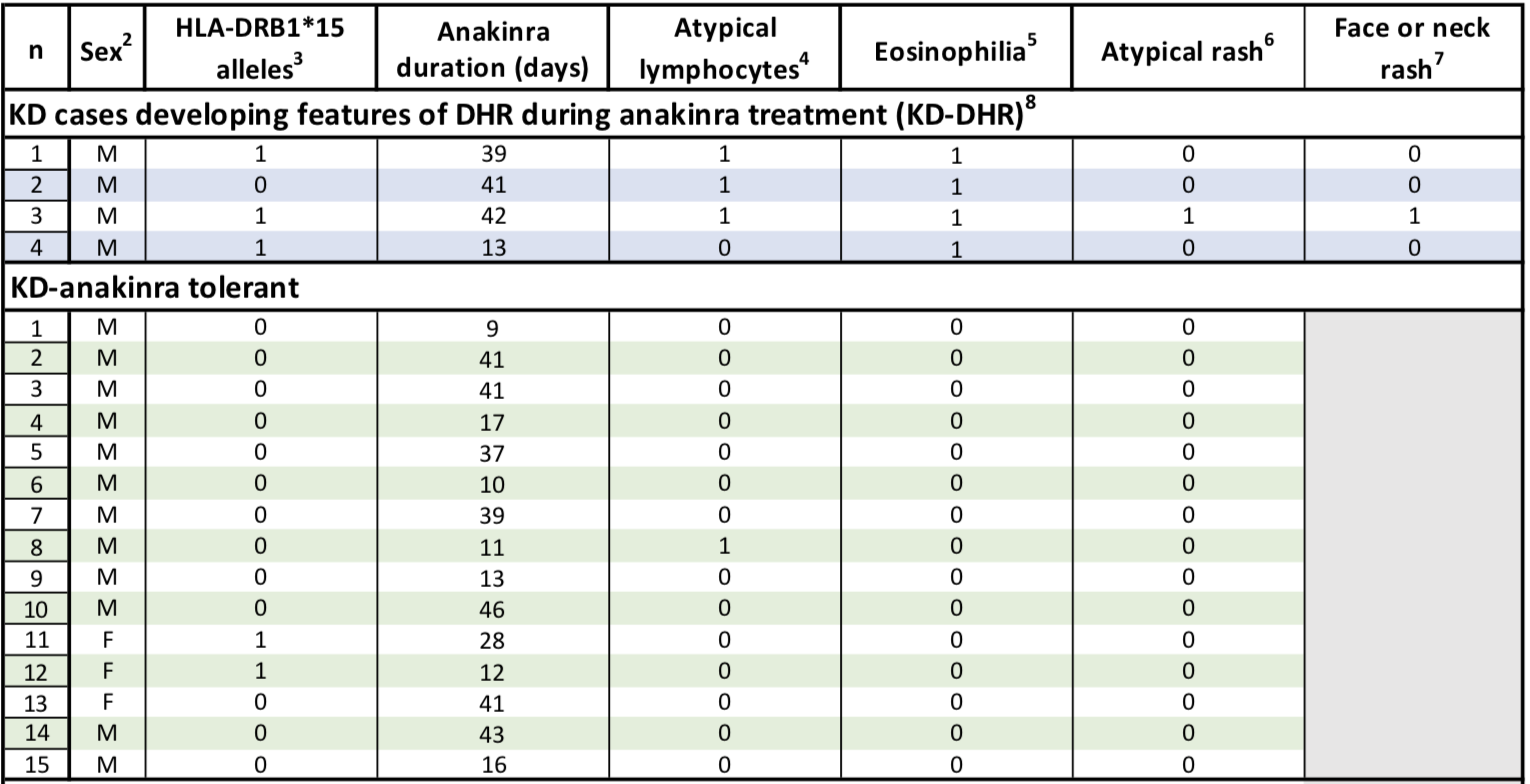

0=absent, 1=present, except for eosinophilia, which is coded as per footnote 5

^1^Clinical features during treatment with anakinra

^2^No significant difference in sex frequency in KD-sAR vs. KD-anakinra tolerant (M vs F: OR 0.5 (95% CI 0.02,11.4), p=0.66)

^3^Presence of HLA-DRB1*15:XX. Cases and controls had different alleles (see table S1b)

^4^Atypical lymphocytes were >2% of WBC and increased >2% from value pre-anakinra.

^5^1= AEC ≥ 500 and elevated over baseline study value (eosinophilic); 2= AEC ≥ 1500/ul and elevated over baseline study value (hyper-eosinophilic). AEC ranged from 630/ul to 1071/ul.

^6^Non-evanescent, intensely pruritic rash, with erythema and localized swelling, beginning during anakinra treatment.

^7^Rash involving the face or neck is highly prevalent (in ~80%) in DRESS-type drug reactions.^26,27^

^8^Data were incomplete for scoring by RegiSCAR. Features scored for DHR were appearance of new rash, increased eosinophilia and atypical lymphocyte counts associated with anakinra treatment.

**Table S4: Association analysis for HLA alleles in sJIA-DHR subjects vs. INCHARGE collection**

| **a. Ancestrally-matched by PCA** | | **Subjects with HLA allele of interest** | | | **sJIA-DHR vs. INCHARGE sJIA** | | **sJIA-DHR vs. INCHARGE controls** | |
| --- | --- | --- | --- | --- | --- | --- | --- | --- |
|  | **Allele** | **sJIA-DHR** | **INCHARGE sJIA** | **INCHARGE controls** | ***P* value** | **OR (95 CI)** | ***P* value** | **OR (95 CI)** |
| **European**  **sJIA-DHR vs. European GWAS subjects** | HLA-DRB1*15:01 | 13/14 (93%) | 130/550 (24%) | 822/3279 (25%) | 4x10^-8^ | 42 (5.4, 324.1) | 7x10^-8^ | 38.9 (5.1, 297.5) |
|  | HLA-DRB1*15:XX | 13/14 (93%) | 136/550 (25%) | 861/3279 (26%) | 8x10^-8^ | 39.6 (5.1, 305) | 1x10^-7^ | 36.5 (4.8, 279) |
|  | HLA-DQB1*06:02 | 13/14 (93%) | 128/550 (23%) | 806/3279 (25%) | 4x10^-8^ | 42.9 (5.6, 330.8) | 5x10^-8^ | 39.9 (5.2, 305.4) |
|  | HLA-DQA1*01:02 | 13/14 (93%) | 188/550 (34%) | 1148/3279 (35%) | 5x10^-6^ | 25 (3.2, 192.8) | 5x10^-6^ | 24.1 (3.2, 184.7) |
|  | HLA-DRB1*11:01 | 4/14 (29%) | 103/550 (19%) | 313/3279 (10%) | NS |  | 0.007 | 7.6 (2.0, 28.4) |
|  | HLA-DQB1*03:01 | 6/14 (43%) | 240/550 (44%) | 1072/3279 (33%) | NS |  | 0.038 | 4.1 (1.0, 16.5) |
| **b. Self-reported "White"** | | **Subjects with HLA allele of interest** | | | **sJIA-DHR vs. INCHARGE sJIA** | | **sJIA-DHR vs. INCHARGE controls** | |
| **"White"**  **sJIA-DHR vs. European GWAS subjects** | HLA-DRB1*15:01 | 24/29 (83%) | 130/550 (24%) | 822/3279 (25%) | 7x10^-11^ | 15.5 (5.8, 41.5) | 8x10^-11^ | 14.3(5.5,37.7) |
|  | HLA-DRB1*15:XX | 24/29 (83%) | 136/550 (25%) | 861/3279 (26%) | 1x10^-10^ | 14.6 (5.5, 39) | 2x10^-10^ | 13.5 (5.1,35.4) |
|  | HLA-DQB1*06:02 | 24/29 (83%) | 128/550 (23%) | 806/3279 (25%) | 6x10^-10^ | 12.6 (5, 31.7) | 7x10^-10^ | 11.8 (4.8, 29) |
|  | HLA-DQA1*01:02 | 23/28 (82%) | 188/550 (34%) | 1148/3279 (35%) | 3x10^-6^ | 7 (2.8, 17.7) | 3x10^-6^ | 6.8 (2.8, 16.8) |
|  | HLA-DRB1*11:01 | 7/29 (24%) | 103/550 (19%) | 313/3279 (10%) | NS |  | 0.019 | 3.2 (1.3, 7.5) |
| **c. All subjects** | | **Subjects with HLA allele of interest** | | |  |  |  |  |
|  | **Allele** | **sJIA-DHR** | **INCHARGE sJIA** | **INCHARGE controls** |  |  |  |  |
| **All sJIA-DHR vs. All GWAS subjects** | HLA-DRB1*15:01 | 39/56 (70%) | 168/773 (22%) | 1649/6812 (24%) |  |  |  |  |
|  | HLA-DRB1*15:XX | 44/56 (79%) | 182/773 (24%) | 1736/6812 (25%) |  |  |  |  |
|  | HLA-DQB1*06:02 | 40/56 (71%) | 171/773 (22%) | 1617/6812 (24%) |  |  |  |  |
|  | HLA-DQA1*01:02 | 43/53 (81%) | 265/773 (34%) | 2322/6812 (34%) |  |  |  |  |
|  | HLA-DRB1*11:01 | 12/56 (21%) | 160/773 (21%) | 649/6812 (10%) |  |  |  |  |

HLA, human leukocyte antigen; sJIA, systemic juvenile idiopathic arthritis; DHR, delayed hypersensitivity reaction; INCHARGE, International Childhood Arthritis Genetics Consortium^5^; GWAS, genome wide association study; PCA, principal components analysis; sJIA-DHR, sJIA with DHR; p-value, p-value by logistic regression; OR (95CI), odds ratio with 95% confidence interval; HLA-DRB1*15:XX, All HLA-DRB1*15 alleles.

HLA-DRB1*15:01~DQA1*01:02~DQB1*06:02 haplotype is in near-complete linkage disequilibrium in European populations.^16^ Analyses of non-White/European subjects from the DHR and INCHARGE cohorts were not performed due to lack of ancestral matching.

**Table S5:** **Published case reports of delayed drug reactions implicating IL-1 or IL-6 inhibitors**

| **Age/Sex** | **Ancestry** | **Condition (inciting drug)** | **Eosinophilia** | **Rash** | **SCAR** | **other organs** | **Outcome** | **Ref** |
| --- | --- | --- | --- | --- | --- | --- | --- | --- |
| 48M | White | rheumatoid arthritis (A) | 1 | 1 | 0 | liver | resolved | 28 |
| adult F | NA | rheumatoid arthritis (T) | 1 | 1 | DRESS | GI, liver | resolved | 29 |
| 52F | NA | rheumatoid arthritis (T) | 1 | 1 | 0 | GI | resolved | 30 |
| 69F | NA | rheumatoid arthritis (T) | 1 | 1 | 0 | none | resolved | 31 |
| 69F | NA | polyarthritis (T) | 1 | 1 | DRESS | HLH | fatal | 32 |
| 55F | NA | adult onset Still’s disease (T) | 1 | 1 | DRESS | liver | resolved | 33 |
| 70M | White | COVID-19 cytokine storm (T) | 1 | 1 | 0 | none | resolved | 34 |
| 82F | Black | giant cell arteritis (T) | NA | 1 | SJS | oral pharynx | resolved | 35 |
| 12F | NA | polyarticular arthritis (T) | 1 | 1 | 0 | fascia | sclerotic skin lesions | 36 |
| 2F | White | autoinflammatory disorder (A) | 1 | 1 | DRESS | liver, HHV6+ | resolved | 37 |
| 2F | Black | autoinflammatory disorder (C) | 1 | 1 | DRESS | lymph nodes | resolved | 37 |
| 2F | White | autoinflammatory disorder (C) | 1 | 1 | DRESS | lung, MAS | improved lung disease | 38 |

1= present, 0= absent; NA, not available; Ancestry, self-identified; A, anakinra; T, tocilizumab; C, canakinumab; SCAR, severe cutaneous adverse reaction; DRESS, Drug reaction with eosinophilia and systemic symptoms; SJS, Stevens-Johnson Syndrome; HLH hemophagocytic lymphohistiocytosis; HHV6+, human herpes virus 6 reactivation; MAS, macrophage activation syndrome

Case reports in conditions other than sJIA and AOSD. DRESS is indicated when reported as the diagnosis. Drug was withdrawn in all cases; outcome ≥ 1 month after drug withdrawal is shown.

48M Lungoci ^28^ , adult F Massolino ^29^ , 52F Morrisroe ^30^, 69F Mori ^31^, 69F Ben Said ^32^, 55F Zuelgaray ^33^, 70M Sernicola ^34^, 82F Venkateswaran ^35^,12F Aeschlimann ^36^

2F anak and 2F cana Polivka ^37^, 2F cana Bader-Meunier ^38^**Supplementary Figures**

**Figure S1**

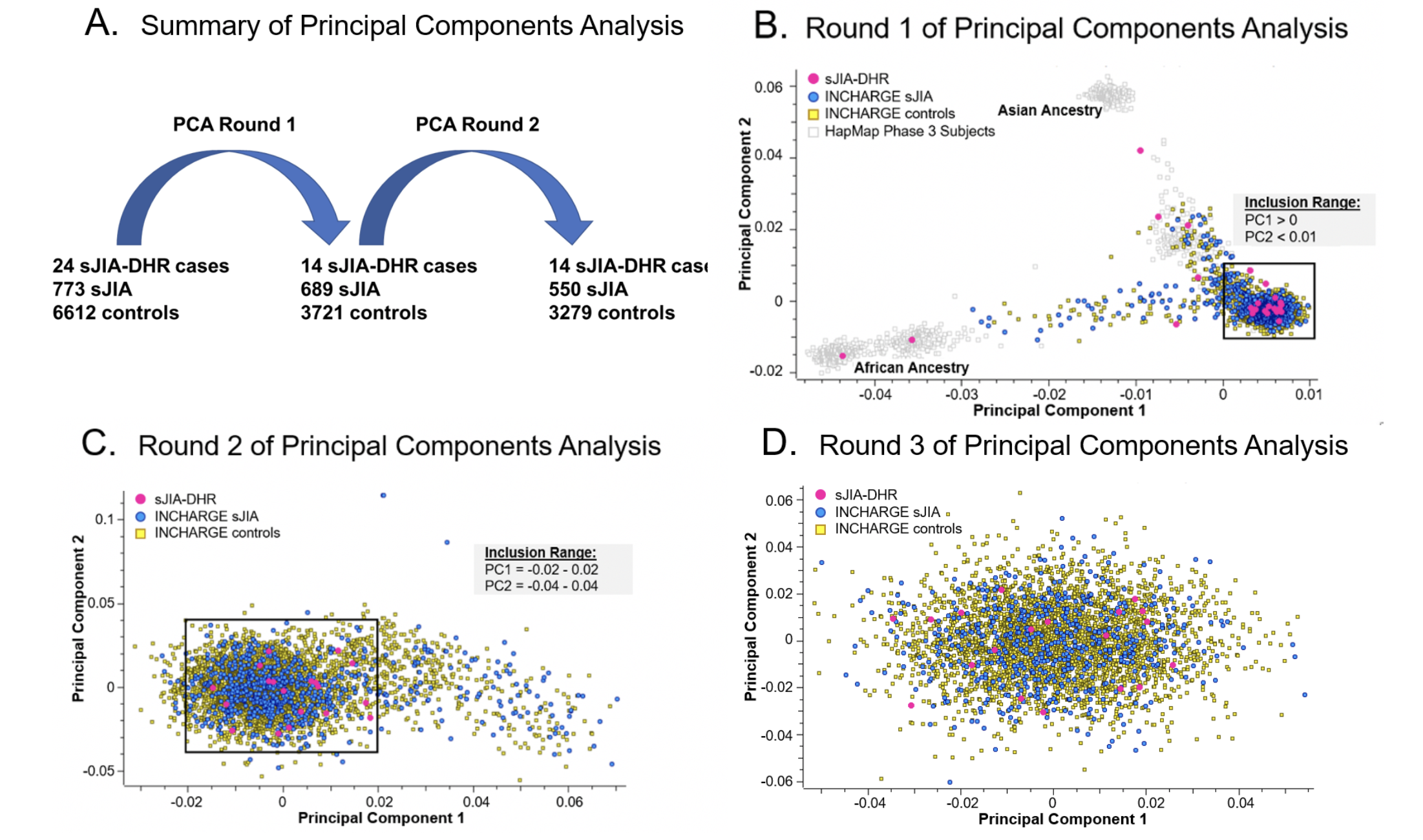

**Supplementary Figure 1. Ancestral matching of INCHARGE sJIA GWAS cases and controls with sJIA-DHR subjects by principal component analysis.** Panel **(A)** demonstrates shows sample accounting through the principal component analysis**. (B)** Principal components (PCs) were calculated using a linkage disequilibrium reduced set of intersecting SNPs between sJIA-DHR subjects (pink circles), INCHARGE sJIA cases (blue circles) and controls (yellow circles), and subjects from the HapMap3^39^ populations (grey boxes). PCs were plotted to identify subjects of European ancestry, marked by the black box, which were carried forward into round 2. **(C)** The second round of PCs were calculated for this reduced set of subjects and were plotted to exclude additional ancestrally dissimilar subjects**. (D)** The third round of PCs were calculated for the subset of subjects defined by round 2, and round 3 PCs were plotted to examine the structure of the final study population. Genomic control inflation factors were calculated for sJIA-DHR vs. sJIA GWAS (λ_GC_=1.042) and sJIA-DHR vs. GWAS controls (λ_GC_=1.056) and indicated robust matching.

Altshuler^25^

Altshuler^39^

**Figure S2**

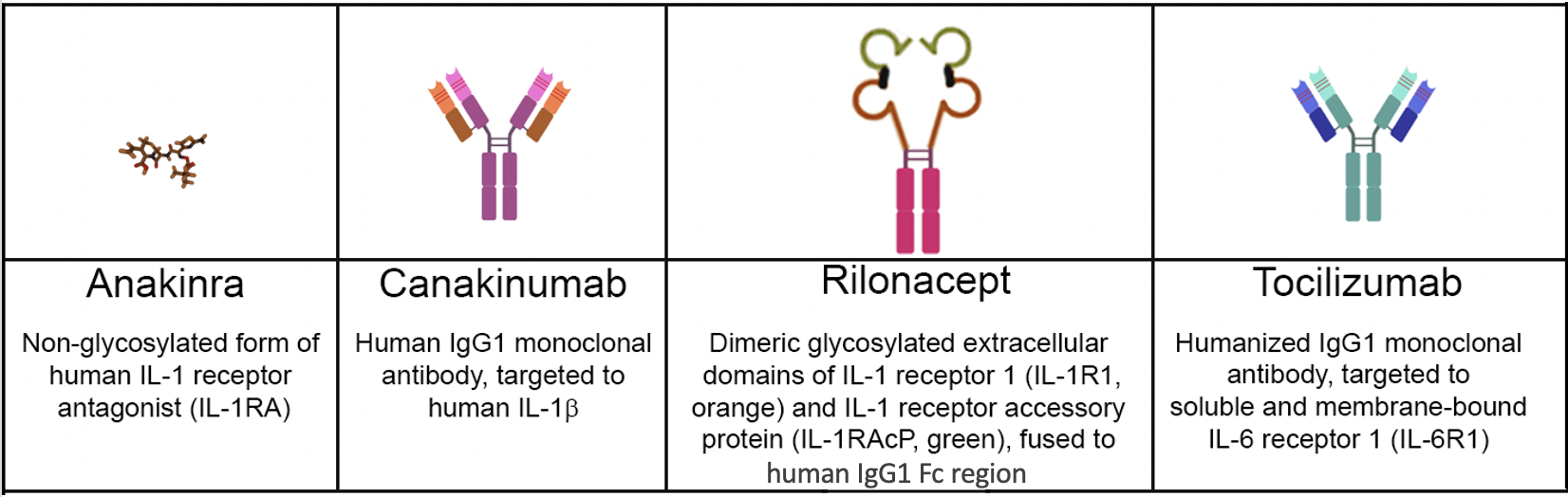

**Supplementary Figure 2. Chemical structures of cytokine inhibitors implicated in HLA-DRB1*15 associated DHR**. Shown are diagrams of the structures of IL-1 inhibitors (anakinra, canakinumab, rilonacept) and an IL-6 inhibitor (tocilizumab), as described in references^40-43^ respectively. The diagrams reflect the relative sizes of the molecules, with the anakinra structure shown at 3X magnification to be visible. Anakinra blocks the biologic activity of IL-1α and IL-1β by competitive inhibition of their binding to IL-1R1. Rilonacept acts as a decoy receptor ("trap") for IL-1β and, with lesser affinity, binds endogenous IL-1RA and IL-1α. Canakinumab binds IL-1β only, and tocilizumab inhibits IL-6 activity by blocking its binding to IL-6R1.A-D Kopf ^40^ 2010,

Kopf ^40^Dhimolea ^41^ 2010 Hoffman ^42^ 2008 Oldfield ^43^2009

6.	The Stanford REDCap platform (<http://redcap.stanford.edu>) is developed and operated by Stanford Medicine Research IT team. The REDCap platform services at Stanford are subsidized by a) Stanford School of Medicine Research Office, and b) the National Center for Research Resources and the National Center for Advancing Translational Sciences, National Institutes of Health, through grant UL1 TR001085.

29.	Massolino RI, Hissaria P, Lee A, Proudman SM. Tocilizumab-induced drug reaction with eosinophilia and systemic symptoms (DRESS) in a patient with rheumatoid arthritis. Rheumatol Adv Pract 2018;2:rky029.
